# The efficacy, effectiveness and safety of SARS-CoV-2 disinfection methods (including ozone machines) in educational settings for children and young people

**DOI:** 10.1101/2022.02.21.22271281

**Authors:** Deborah Edwards, Judit Csontos, Elizabeth Gillen, Ruth Lewis, Alison Cooper, Micaela Gal, Rebecca-Jane Law, Adrian Edwards

## Abstract

While evidence for the importance of transmission of SARS-CoV-2 from contaminated surfaces is limited, ozone disinfection methods have been considered for surface cleaning as a response to stopping the spread of the virus in educational settings. This rapid evidence summary aimed to search the available literature and summarise findings on the surface survival of SARS-CoV-2, efficacy and effectiveness of ozone machines against SARS-CoV-2, and benefits and harms caused by using these cleaning technologies, including their impact on health. Alternative cleaning technologies, such as light-based technologies and hydrogen peroxide vapour, were also investigated. Findings indicate that gaseous ozone can inactivate different bacteria and viruses, although there is a lack of direct evidence investigating the effect of these cleaning methods on SARS-CoV-2 in real-world settings, specifically in schools. However, regarding harm, ozone is a highly reactive oxidising agent, and high concentrations can contribute to decay of building materials, and health issues (mainly respiratory) by direct exposure or by-product formation. Therefore, leading environmental health organisations do not recommend the use of ozone cleaning technologies in real-world settings, such as schools. Research and policy focus may need to shift towards other interventions that could help reduce transmission, and consequently minimise disruption to education.

**Funding statement:** The Wales Centre for Evidence Based Care was funded for this work by the Wales Covid-19 Evidence Centre, itself funded by Health & Care Research Wales on behalf of Welsh Government.

**TOPLINE SUMMARY:** *What is a Rapid Evidence Summary?:* This Rapid Evidence Summary was completed in two weeks to inform policy-decision making. It is based on a systematic search of the literature, conducted in September 2021. Priority is given to studies representing robust evidence synthesis. No quality appraisal or evidence synthesis are conducted, and the summary should be interpreted with caution.

*Background / Aim of Rapid Evidence Summary:* Several non-touch disinfectant methods including ozone, light-based technologies, and hydrogen peroxide are being considered to reduce the risk of SARS-CoV-2 virus transmission to children and young people in educational settings. Concerns have been raised about the evidence of efficacy, effectiveness and safety of these technologies in these settings. We aimed to address the following research questions:

- What is the evidence for the **surface survival of SARS-CoV-2?**
- What is the evidence for the efficacy (in vitro) and real-life effectiveness (in situ) of ozone machines, light-based technologies and hydrogen peroxide vapour as air or surface disinfectants against SARS-CoV-2?
- What are the potential health effects of ozone, in particular for children and young people and the benefits and harms of using ozone machines?

*Key Findings:* Extent of the evidence base
A total of **82** tertiary, secondary and primary evidence sources was included Recency of the evidence base
Most studies were published **2020-21**, indirect evidence was included from earlier work from 2006 onwards Summary of findings

- SARS-CoV-2 fragments can be found on surfaces **up to seven days** later in the community but there is a lack of evidence whether these are viable
- When accounting for both surface survival data and real-world transmission factors, the risk of surface transmission after a person with COVID-19 has been in an indoor space is minor after 72 hours, regardless of last clean
- There is evidence from **experimental settings** that ozone machines, light-based technologies and hydrogen peroxide do inactivate coronaviruses, including SARS-CoV-2
- There is a lack of evidence for the effectiveness of ozone machines, light-based technologies and hydrogen peroxide in real-world settings
- There are **uncertainties** about training requirements for staff, methods for assurance of ozone removal and monitoring of occupational exposure
- There is strong evidence of a **causal relationship between short term ozone exposure and respiratory health issues;** these can occur at very low concentrations of ozone; children with asthma are more at risk
- Rooms using ozone machines need to be **sealed off to avoid leakage** of the ozone gas which is **toxic at high concentrations**
- Ozone may react with materials in the room to form **secondary pollutants** (e.g. formaldehyde) The best quality evidence

- The <u>US EPA 2021</u> does not recommend ozone for air cleaning and the <u>UK SAGE EMG</u> 2020a does not recommend technologies that “may generate undesirable secondary chemical products that could lead to health effects such as respiratory or skin irritation (medium confidence). These devices are therefore not recommended unless their safety and efficacy can be unequivocally and scientifically demonstrated by relevant test data” (SAGE EMG 2020a).

*Policy implications:*

- There is **no direct evidence for the effectiveness and safety** of using ozone machines to deactivate SARS-CoV-2 **in real-world educational settings** for children, young people and staff
- There is evidence for the **risk of potential harm** to children and young people of ozone machines from either ozone or secondary pollutants, in particular but not only, if **used in uncontrolled ways in educational settings**

*Strength of Evidence to date:*

- moderate evidence for the surface survival of SARS-CoV-2
- strong evidence of causal relationship between short term ozone exposure and respiratory health issues

## 1. Context / Background

It is possible for people to be infected with SARS-CoV-2 through contact with contaminated surfaces or objects (fomites), but the risk is generally considered to be low (CDC 2021). For surface transmission to occur, enough viable virus must reach the susceptible host following transfer to a surface, survival on the surface, transfer from the surface to skin, and from the skin to mucous membrane. Effective hand hygiene disrupts both surface transmission and direct contact, and routine cleaning and disinfection helps address any remaining risks of SARS-CoV-2 infection arising from contact with contaminated surfaces. Additional cleaning in response to an outbreak within an educational setting may be desirable. A number of automated room disinfection systems have been reported across the literature and include: the use of **aerosolized and vapoured hydrogen peroxide** (Choi et al 2020; Dancer 2014; Rutala and Weber 2013; SAGE 2020a; Tarka and Nitsch-Ousch 2020, Tsou et al. 2020; Weber et al. 2016; Weber et al 2019), **chlorine dioxide vapour** (Davies et al 20211; Otter et al 2020; Rutala and Weber 2013; Tarka and Nitsch-Ousch 2020; Tsou et al. 2020**), light-based technologies** (Boyce 2016; Choi et al 2020; Otter et al 2020; Dancer 2014; SAGE 2020a; Tarka and Nitsch-Ousch 2020, Tsou et al. 2020; Weber et al. 2016. Weber et al. 2019) and **ozone** (Dancer 2014; Otter et al. 2013; Tarka and Nitsch-Ousch 2020, Tsou et al. 2020).

We are aware that ozone disinfectant machines are being used in a school in Dublin (see <u>Dublin school credits ozone disinfecting machines for preventing Covid outbreaks - Dublin Live</u>). However, the Chair of the Irish Government’s Expert Group on the Role of Ventilation in Reducing Transmission of COVID-19 has confirmed to us that to the best of his knowledge (24^th^ September 2021), there has been no evaluation process undertaken by the Government of Ireland to address the safety, efficacy, and feasibility of ozone disinfection devices in educational settings. Discussions in Ireland continue in the context that there do not appear to have been any real-world studies which have investigated its efficacy, and that the use of ozone presents a number of major drawbacks in relation to toxicity (at high concentrations, ozone inhalation can damage the lungs and exacerbate asthma), reaction with organic compounds in air and on surfaces to produce potentially hazardous air pollutants (e.g. formaldehyde and ultra-fine particles) and at high concentrations, ozone can also damage materials such as rubber, fabric, and electrical wire coatings. The Irish Health and Safety Authority (HSA) has very recently updated their guidance for returning to the workplace: https://www.gov.ie/en/publication/22829a-return-to-work-safely-protocol/. Under the section Other Equipment and Systems (page 49), there is a new addition in relation to ozone machines: “Other devices such as ozone generating devices and air disinfection devices may present additional chemical related hazards in the workplace and their use should be fully justified by an appropriate risk assessment. It is not recommended to use these devices in occupied spaces.”

## 2. Research question(s)

Q1: What is the evidence for the **surface survival of SARS-CoV-2?**

Q2a: What is the evidence for the potential efficacy of ozone as an air or surface disinfectant against SARS-COV-2?

Q2b What is the evidence for the potential efficacy of light-based technologies as air or surface disinfectants against SARS-CoV-2?

Q2c What is the evidence for the potential efficacy of hydrogen peroxide as an air or surface disinfectant against SARS-CoV-2?

**Q3a**: **What is the** effectiveness of no-touch automated ozone disinfection methods for decontamination against SARS CoV-2?

Q3b: What is the effectiveness of no-touch automated light-based disinfection methods for decontamination against SARS CoV-2?

Q3c: What is the effectiveness of no-touch automated aerosolized or vapoured hydrogen peroxide disinfection methods *for decontamination against SARS CoV-2?*

Q4a: What are the potential health effects of ozone for children and young people?

Q4b: What are the **benefits** and **harms** of using **ozone machines?**

## 3. Summary of the evidence base

### 3.1 Type and amount of evidence available

**Q1: What is the evidence for the surface survival of *SARS-CoV-2*?**

- **Two rapid reviews** (NCCMT 2021, NHLKS 2020), **three systematic reviews** (Bedrosian et al. 2021, Kampf et al. 2020a; Marzoli et al. 2021) and **one science brief** (CDC 2021) investigated the surface survival of SARS-CoV-2.
- The systematic review conducted by Bedrosian et al. 2021 was also included in the rapid review conducted by the NCCMT 2021 and the systematic review by Kampf et al. 2020a was included in the rapid review conducted by NHLKS 2020.

**Q2a: What is the evidence for the potential efficacy of <u>ozone</u> as an air or surface disinfectant against *SARS-CoV-2*?**

- **One systematic review** (Bayarri et al 2021) and **six narrative reviews** (Alimohammadi and Naderia 2021; Blanco et al 2021; Davies et al 2011; Grignani et al. 2021; Morrison et al. 2021; Otter et al. 2013) investigated the ability of **ozone** to inactivate a variety of bacteria and viruses including **SARS-CoV-2 surrogates.**
- The use of automated room disinfection systems in test environments using ozone was investigated in **six primary studies** (Dubuis et al 2020; Franke et al 2021; Hudson et al. 2007; Knobling et al 2021; Steinmann et al 2021; Uppal et al 2021).

**Q2b: What is the evidence for the potential efficacy of <u>light-based technologies</u> as air or surface disinfectants against *SARS-CoV-2*?**

- **Three systematic reviews** (Chiappa et al. 2021; Kwok et al. 2021; Ramos et al 2020), **four narrative reviews** (Hadi et al. 2020; Hessling et al 2021; Memarzadeh 2021; Raeiszadeh and Adeli 2020) and **one protocol for a systematic review** (de Oliveira et al. 2020) investigated the efficacy of **ultraviolet technologies** against **coronaviruses.**
- Additionally, two **SAGE EMG reports** and **one TAG-E** explore the potential of light-based technologies against SARS-CoV-2 in terms of air cleaning (SAGE-EMG 2020a; SAGE-EMG 2020, TAG-E 2021).

***Q2c: What is the evidence for the potential efficacy of <u>hydrogen peroxide</u> as an air or surface disinfectant against coronaviruses?***

- **One systematic review** (Kampf et al 2020a, b), **one rapid review** (Shimabukuro et al. 2020); **one letter to the editor** (Lopez Ortega et al. 2020), **one protocol for a systematic review** (de Oliveira et al. (2020) explored the efficacy of **hydrogen peroxide** as an air and surface disinfectant against **coronaviruses.**
- Additionally, **two SAGE EMG reports** and **one TAG-E** explore the potential of hydrogen peroxide vapour against SARSCoV-2 in terms of air cleaning (SAGE-EMG 2020a; SAGE-EMG 2020, TAG-E 2021).

***Q3a: What is the* effectiveness of no-touch automated ozone disinfection methods *for decontamination against SARS CoV-2?***

- **One primary study** assessed the inactivation of airborne and surface contaminants in healthcare structures using ozone (Moccia et al. 2020).
- **One rapid review** looked for data for the effectiveness of no-touch modalities (including ozone) for disinfecting patient rooms in hospital or acute care settings for respiratory viral pathogens and other pathogens with potential relevance to assessing effectiveness in SARS-CoV-2 (Tsou et al. 2020).
- **One narrative review** looked for data for the effectiveness of no-touch modalities (including ozone) for disinfecting dental clinics during the Covid-19 pandemic (Scarano et al. 2020).

***Q3b: What is the *effectiveness* of no-touch automated light-based disinfection methods for decontamination against SARS CoV-2?***

- **One systematic review** (Alvarado-Miranda et al 2020) investigated the effectiveness of UV light technologies for disinfection of contaminated spaces in healthcare settings.
- **One rapid review** looked for data for the effectiveness of no-touch modalities (including ultraviolet light) for disinfecting patient rooms in hospital or acute care settings for respiratory viral pathogens and other pathogens with potential relevance to assessing effectiveness in SARS-CoV-2 (Tsou et al. 2020).
- **One narrative review** looked for data for the effectiveness of no-touch modalities (including ultraviolet light and Pulsed xenon) for disinfecting dental clinics during the Covid-19 pandemic (Scarano et al. 2020).
- Two protocols for systematic reviews have been published that intend to explore the effectiveness of a variety of light-based technologies for disinfection of surfaces contaminated with SARS-CoV-2.
  - light-based therapies **(Biophotonics**) as antiviral therapy, as well as anti-coronavirus therapy (Cecatto et al. 2020).
  - **UV-C light technology** in dental and medical practices for surface disinfection (de Melo-Monteiro 2020).

***Q3c: What is the effectiveness of no-touch automated aerosolized or vapoured hydrogen peroxide disinfection methods for decontamination against SARS CoV-2?***

- **One rapid review** looked for data for the effectiveness of no-touch modalities (including hydrogen peroxide vapour) for disinfecting patient rooms in hospital or acute care settings for respiratory viral pathogens and other pathogens with potential relevance to assessing effectiveness in SARS-CoV-2 (Tsou et al. 2020).
- **One narrative review** looked for data for the effectiveness of no-touch modalities (including aerosolized hydrogen peroxide and H2O2 vapour) for disinfecting dental clinics during the Covid-19 pandemic (Scarano et al. 2020).

***Q4a: What are the potential health effects of ozone for children and young people***

- **Three systematic reviews** (Atkinson et al. 2016; Gao et al. 2020; Huangfu and Atkinson 2020), **one evidence synthesis** based on systematic review methodology (US EPA 2020) and **one protocol for a systematic review** (Zhao e al. 2020) investigated the health effects of either short term and/or long-term exposure to ozone.
- A further **narrative review** commented on the health effects regarding exposure to ozone in indoor settings (Namdari. et al. 2021).
- Additionally, the **SAGE EMG report from** November 2020 that explored the potential application of air cleaning devices and personal decontamination to manage transmission of COVID-19 has a section on the potential health effects of air cleaning devices (SAGE-EMG 2020a).
- The **US EPA 2021** have produced a **web-based summary** on ozone generators that are sold as air cleaners that also contributed to the evidence base.

***Q4b: What are the benefits and harms of using ozone machines?***

| Review question |  |
| --- | --- |
| **What are the efficacy (in vitro), effectiveness (in situ) and safety of SARS CoV-2 disinfection methods (including ozone machines) in educational settings for children and young people?** |  |
| **Participants** | Children, young people and adults potentially coming into contact with SARS-CoV-2 in any indoor setting outside of the home environment |
| **Intervention** | Disinfection methods (including ozone machines) |
| **Comparison** | Any |
| **Outcomes** | Inactivation of SARS-Cov-2 or adverse effects |
| **Other Study Considerations** |  |
| Other viruses and bacteria were also considered where evidence was lacking for SARS-Cov-2 |  |

Information from within the results or the discussion of **seven narrative reviews** (Alimohammadi M and Naderia M. 2021; Blanco et al. 2021; Davies et al. 2021; Grignani et al. 2021; Otter et al. 2013; Moccia et al. 2020; Morrison et al. 2021; Namdari et al. 2021), **five primary studies** (Franke et al. 2021; Hudson et al. 2007; Knobling et al. 2021) and **eight secondary sources** ((Druzik 1985; Kamaruddin and Muhr 2018; Lee et al 1996; Morrison and Nazaroff 2002; Morrison et al. 1998; Poppendieck et al. 2007a, Poppendieck et al. 2007b; Weschler 2000) provided information on the benefits and/or harms of using ozone machines.

The US EPA 2021 have produced a **web based summary** on ozone generators that are sold as air cleaners that also contributed to the evidence base.

### 3.2 Key Findings

***Q1: What is the evidence for the surface survival of SARS-CoV-2?***

- Out of the three reviews reported, two looked at studies and evidence syntheses investigating surface survival in vitro and in situ (NCCMT 2021, NHLKS 2020), while Marzoli et al. (2021) only looked at in vitro primary research.
- The NHLKS (2020) report was conducted relatively early in the SARS-CoV-2 pandemic, thus their main source of information was single studies on SARS-CoV-2, guidelines, expert opinion, and reviews of the SARS and MERS family.
- There is **moderate GRADE evidence that SARS-CoV-2 fragments can be found on surfaces up to seven days** in the community. However, there is a lack of evidence whether these viral fragments are viable (have the potential to cause infection) (NCCMT 2021).
- NHLKS (2020) reported that viable SARS-CoV-2 can be found on surfaces for up to 72 hours, although virus titre reduced over time.
- When accounting for both surface survival data and real-world transmission factors, the **risk of fomite transmission** after a person with COVID-19 has been in an indoor space is **minor after 3 days** (72 hours), regardless of when it was last cleaned (CDC, 2021).
- Virus survival was greatly dependant on surface composition, temperature, and humidity (NCCMT 2021, NHLKS 2020, Marzoli et al. 2021).
- Viable virus could be detected longer on smoother surfaces, such as stainless steel and plastic (NCCMT 2021)
- Marzoli et al. (2021) also reported glass, and paper banknotes as surfaces that were optimal for virus survival up to 28 days.
- Materials on which viable virus survived for a shorter period included cardboard, cotton, and copper (NCCMT 2021, NHLKS 2020).
- There was a consensus between reviews, that SARS-CoV-2 had more rapid decay with increasing temperature and humidity (CDC, 2021; NCCMT 2021, NHLKS 2020, Marzoli et al. 2021).
- As to what virus survival on surfaces means regarding transmission, there is low GRADE evidence that SARS-CoV-2 fragments on surfaces may not be viable (NCCMT 2021).

**Summary in vitro survival** - SARS-CoV-2 could survive for between **3 to 28 days** depending on the type of surface, and the initial virus titre placed on the surface.

**Summary in situ survival** - SARS-CoV-2 fragments can be found on surfaces for between **3 to 7 days** in the community.

***Q2a: What is the evidence for the potential efficacy of <u>ozone</u> as an air or surface disinfectant against SARS-Cov-2?***

- Ozone has been shown to be **effective in inactivating** a wide variety of bacteria and viruses (such as Norovirus feline calicivirus, influenza) including **SARS-CoV-2 surrogates** (other coronaviruses and enveloped viruses).
- Ozone disinfection systems are more effective when used in combination with high humidity (Bayarri et al 2021; Dubuis et al. 2020; Franke et al. 2021).
- All the studies reported that at the end of the process the active substance was completely degraded and that there was no residual ozone or measurable increases in ozone detected.
- One study reported that levels of ozone in the room are continuously displayed on a mobile tablet computer and recorded in a standardized manner (Knobling et al. 2021).

***Q2b: What is the evidence for the potential efficacy of <u>light-based technologies</u> as an air or surface disinfectant against coronaviruses?***

- Ultraviolet light technologies (UVA, UVB and UVC) have been shown to be **effective in inactivating** coronaviruses and have high viricidal potential against **coronaviruses.**
- There is **good evidence** that germicidal UV (GUV) that uses UV-C light is likely to be **viable decontamination approach** against SARS-CoV-2 for unoccupied rooms (SAGE-EMG 2020a; SAGE 2020b, TAG-E, 2020).
- **Complete inactivation** of coronaviruses on surfaces or aerosolized, including SARS-CoV-2, was reported to take a maximum **exposure time of 15 min** and to need a maximum distance from the **UV emitter of up to 1 m (**Chiappa et al 2021).
- **Coronaviruses** are very **UV sensitive** Since coronaviruses do not differ structurally to any great extent, the SARS-CoV-2 virus – as well as possible future mutations – will very likely be highly UV sensitive, (Hessling et al 2020).
- The efficacy of light-based inactivation was **reduced by the presence of absorptive materials** (Hadi et al 2020).
- Experimental studies on ssRNA viruses, even the early strains of the CoV family, strongly support the opinion that the **SARS-CoV-2 can be inactivated by UV radiation** (Raeiszadeh and Adeli 2020).
- The **germicidal effect of UV-C** is potent against microorganisms including viruses, methicillin-resistant Staphylococcus aureus, and vancomycin-resistant enterococci however, **direct evidence is needed** for any targeted implementation of UV-C during **COVID-19 pandemic** (Ramos et al 2020).
- Different experimental methods testing UV light have shown that it can **inactivate the Coronaviruses,** including SARS-CoV-2, SARS-CoV-1, MERS-CoV virus (Kwok et al. 2021).
- The efficacy of UV-based sanitizer technologies are promising but are dependent on numerous environmental, physical and technical factors (Memarzadeh 2021).

***Q2b: What is the evidence for the potential efficacy of <u>hydrogen peroxide</u> as an air or surface disinfectant against coronaviruses?***

- **Coronaviruses** can be **efficiently inactivated** by hydrogen peroxide in experimental and hospital settings (de Oliveria et al. 2020; Kampf et al 2020a, b; Shimabukuro et al. 2020)
- There is **good evidence** that fumigation approaches (particularly Hydrogen Peroxide Vapour) are **likely to be viable** decontamination approaches **against SARS-CoV-2** for unoccupied rooms (SAGE-EMG 2020a).

***Q3a: What is the effectiveness of no-touch automated ozone disinfection methods for decontamination against SARS CoV-2?***

- One study was found that was conducted in situ in a healthcare setting and demonstrated **a significant reduction in the microbial count** that always fell below the threshold value after decontamination with an **ozone generator** (Moccia et al. 2020).
- One rapid review **did not find any studies for ozone as a method of disinfection** for respiratory pathogens in acute settings and so effectiveness remains unclear (Tsou et al. 2020).
- One narrative review **did not find any studies** for **ozone as a method of disinfection** for disinfecting dental clinics during the Covid-19 pandemic (Scarano et al. 2020).
- The <u>US EPA 2021</u> does not recommend ozone for air cleaning and the <u>UK SAGE EMG</u> 2020a does not recommend technologies that “may generate undesirable secondary chemical products that could lead to health effects such as respiratory or skin irritation (medium confidence). These devices are therefore not recommended unless their safety and efficacy can be unequivocally and scientifically demonstrated by relevant test data” (SAGE EMG 2020a).

***Q3b: What is the effectiveness of no-touch automated light-based disinfection methods for decontamination against SARS CoV-2?***

- The rapid review by Tsou et al. 2020 only retrieved one systematic review that contained three studies relating to ultraviolet irradiation as a method of disinfection for respiratory pathogens in acute settings and so the effectiveness is unclear (Tsou et al. 2020).
- One narrative review **did not find any studies** for **Ultraviolet C light or pulsed xenon as** a method of disinfection for disinfecting dental clinics during the Covid-19 pandemic (Scarano et al. 2020).
- **40% of the studies** within the review by Alvarado-Miranda et al. 2020 **did not find sufficient scientific evidence** to determine the effectiveness of UV technology on the control of the spread of pathogens in infected areas.

***Q3c: What is the effectiveness of no-touch automated aerosolized or vapoured hydrogen peroxide disinfection methods for decontamination against SARS CoV-2?***

- The rapid review by Tsou et al. 2020 only retrieved one systematic review that contained one study relating to hydrogen peroxide vapour as a method of disinfection for respiratory pathogens in acute settings and so the effectiveness is unclear (Tsou et al. 2020).
- One narrative review **did not find any studies** for **aerosolized hydrogen peroxide and H2O2 vapour as** a method of disinfection for disinfecting dental clinics during the Covid-19 pandemic (Scarano et al. 2020).

***Q4a: What are the potential health effects of ozone for children and young people***

The effects of ozone on health are widely reported and include:

- The UK Committee on the Medical Effects of Air Pollutants (COMEAP) does not consider there to be a safe level of exposure.
- When inhaled at high concentrations, ozone can damage the lungs and exacerbate asthma (US EPA 2020, USA EPA 2021).
- Respiratory irritation (e.g., cough, shortness of breath) even at low levels (SAGE- EMG 2020a; Namdari et al. 2021, US EPA 2021).
- Early findings from Zhao et al. (2020), regarding COPD and reported in a conference abstract, indicate that the results of epidemiological studies are mixed and inconclusive. However, human exposure studies reported that inhalation of ozone caused lung dysfunction and airway hyperresponsiveness
- Chest pain, respiratory inflammation, airway tissue damage, a decrease in lung function, deadening the sense of smell, eye irritation, headache, and exacerbation of respiratory diseases and cardiovascular problems (Namdari et al. 2021, US EPA 2021).
- Findings on long-term exposure to ozone and mortality were equivocal. Huangfu and Atkinson (2020) found an association with all cause and respiratory mortality, whereas an earlier review, Atkinson et al. (2016), had concluded that there was no association with risk of death from all causes, cardiovascular or respiratory diseases or lung cancer. Both reviews on long-term Ozone exposure noted limitations due to the paucity of evidence and substantial heterogeneity within the evidence base.
- Short-term exposure to ambient ozone was associated with increased risk of COPD hospitalisations (Gao et al 2020).
- Recovery from the harmful effects can occur following short-term exposure to low levels of ozone, but health effects may become more damaging and recovery less certain at higher levels or from longer exposures (US EPA 2021)
- Available scientific evidence shows that at concentrations that do not exceed public health standards, ozone has little potential to remove indoor air contaminants, viruses, bacteria, mould or other biological pollutants (US EPA 2021).
- Ozone concentrations would have to be 5-10 times higher than public health standards allow before the ozone could decontaminate the air sufficiently to prevent survival and regeneration of the organisms once the ozone is removed (US EPA 2021).
- Formaldehyde, a by-product (**secondary pollutant**) of the ozone disinfection process, also causes respiratory irritation even at low levels and is harmful to exposed mucous membranes. This chemical is also carcinogenic (SAGE-EMG 2020a).

A summary of the United States Environmental Protection Agency (EPA) (2020) Integrated Science Assessment (ISA) for Ozone and Related Photochemical Oxidants evidence synthesis and its detrimental effects on health is provided below:

#### General overview

- Gaseous ozone can be found in the Earth’s upper atmosphere and at ground level. However, ground level ozone, which is a key element of urban smog, can contribute to potential health issues.
- Inhalation of ozone can react with the epithelial lining of the respiratory system, particularly with lipids, proteins, and antioxidants.
- There is evidence that exposure to ozone can result in autonomic, endocrine, immune, and inflammatory reactions throughout the body, thus potentially leading to respiratory, metabolic, and cardiovascular health problems.

*There is **strong evidence** base for a causal **relationship between short-term ozone exposure and respiratory health issues***.

- Lung function of healthy adults can be affected at concentrations as low as 60 parts per billion (ppb), and respiratory symptoms can occur at 70 ppb or over following 6.6 hours of exposure during exercise. The current US maximum limit of ambient air concentration is 70ppb.
- Ozone can also induce respiratory inflammations at 60 ppb with a 6.6-hour exposure. Furthermore, epidemiological studies in the USA show that ozone exposure has a causal relationship with emergency department (ED) visits for asthma exacerbations.
- The relationship between ED visits and ozone exposure was highest among children between ages 5 and 18, indicating that children with asthma are a high-risk population. The levels of ozone leading to ED admission ranged between 31 and 54 ppb for an average 8 hours.
- Not only children with asthma, but children in general are an at-risk population regarding ozone exposure, as the human respiratory system is in development until age 18 and 20, and toxic gases can lead to issues with normal lung evolution.
- Long-term exposure to ozone has a likely causal relationship with new onset asthma in children with an average annual ozone concentration of 32.1 ppb (ranging from 26 to 76 ppb) based on epidemiological studies.

#### Regarding the general population

- There is a possible relationship between long-term ozone exposure and COPD hospitalisation with mean levels of ozone at 39.3 ppb.
- There is a likely causal relationship between short-term exposure to ozone and metabolic diseases, such as higher level of triglycerides, fasting hyperglycaemia, low HDL, high blood pressure, and central adiposity, although these findings are mainly based on animal studies.
- There is epidemiological evidence for association between diabetic ketoacidosis and coma, and short-term ozone exposure (mean ozone concentration of 64.4 ppb over 24 hours) in older adults (over 75 years).

There is **some evidence** for **other health effects of ozone exposure**

- Such as increased mortality, cardiovascular, nervous, and reproductive system issues.
- Evidence supporting these associations are much weaker, and scarce, mainly relying on animal toxicity studies.

#### Details of the evidence base

- The findings of this EPA (2020) review are based on epidemiological, controlled healthy adult human exposure, and animal toxicity studies.
- EPA screened 309 studies on the HERO database with regards to respiratory issues, 189 on cardiovascular issues, 89 on metabolic health problems, 72 on mortality, and 411 on other health issues.
- Based on EPA’s (2020) description, the quality of papers included in this report was systematically assessed with a focus on the strengths, limitations, and uncertainties of the overall evidence, leading to the description of the level of causality.
- Potential issues with the evidence base included comparability of animal toxicity and controlled human exposure studies.
- Animal toxicity studies often used higher ozone concentration than the general ground level ozone, or what was acceptable in controlled human exposure studies.
- The controlled human exposure studies involved healthy participants, while animal toxicity studies were conducted in a rodent model of disease states. Other differences between animal and human exposure studies included timing of exposure, and temperature. Issues regarding the epidemiological studies included exposure measurement errors, which could potentially lead to bias.

***Q4b: What are the benefits and harms of using ozone machines?***

#### Potential benefits of using of ozone machines in educational settings

- As it is a gas it can **penetrate every part of the room** (Blanco et al. 2021; Hudson et al. 2007).
- Gas is **easy** and economical to **produce** (Hudson et al. 2007).
- **Decays quickly** back to oxygen with half-life of about 20 minutes (Hudson et al. 2007).
- The use of a **catalytic converter** (scrubber) considerably **speeds up the removal** of the gas (Hudson et al. 2007).
- For better protection, **ozone destructors** can be used and operated in the hallway near the closed door of the hospital rooms and inside them when the treatment is completed (Dubuis et al. 2020).
- The gas is readily **detected by smell** and hence can be avoided (Hudson et al. 2007) but it has been noted that over time ozone will quickly damage the ability of a person to smell.

#### Potential harms of using ozone machines in educational settings

- **Long processing times** dependent on the individual machine (Knobling et al. 2021).
- **Toxicity at high concentrations** (above 0.1ppm - Dubuis et al. 2020; Otter et al. 2013) so **rooms (**doors, windows and ventilation diffusers) **need to be sealed** off or quarantined for the duration of the treatment so that effective containment of the gas can be achieved (Alimohammadi and Naderia 2021; Dubuis et al 2020; Franke et al. 2007; Hudson et al. 2007; Knobling et al. 2021; Moccia et al. 2020; Otter et al. 2013) and **monitoring for leakage** and measurements to assure that the room is safe to enter on completion (Otter et al. 2013).
- Due to the generated water aerosol, **smoke detectors must also be covered** to avoid unwanted alarms (Franke et al. 2021; Knobling et al. 2021).
- **Do not use** in the presence of **flammable substances** such as alcohol, petrol, hydrocarbons, bromine, hydrobromic acid, nitrogen oxides and nitroglycerin (Moccia et al 2020).
- **Avoid exposure to UV rays** produced by fluorescent lamps (Moccia et al. 2020).
- **Highly reactive and toxic gas** which has the potential for:

`o` **The decay of building materials** -rubber or derived composites products, surface coatings (Alimohammadi and Naderia 2021; Blanco et al. 2021; Davies et al. 2011; Dubuis et al. 2020; Knobling et al. 2021; Grignani et al. 2021; Morrison et al. 2021; Otter et al. 2013. secondary sources cited Kamaruddin and Muhr 2018; Lee et al 1996).
`o` Additionally, ozone may **react with a range of materials** commonly anticipated in the **indoor environment including** paint, linoleum, carpet, paper, wood and semi-volatile organics adsorbed to surfaces (Morrison et al. 2021; secondary sources cited Weschler 2000).
`o` **Fading dyes on nylon** and acetate and most of the natural dyes and dye-based pigments used by artists (Grignani et al. 2021; Morrison et al. 2021; secondary sources cited Druzik 1985).
`o` **Corrosion of metals** such as copper and aluminium (Davies et al. 2011; Grignani et al. 2021, US EPA 2021); secondary sources cited Druzik 1985).
- Ozone takes part in **reactions with organic pollutants** present in the indoor environment:

`o` Subsequent **formation of secondary pollutants** (Dubuis et al. 2020; Knobling et al. 2021; Grignani et al. 2021; Morrison et al. 2021; secondary sources cited Morrison and Nazaroff 2002; Morrison et al. 1998; Poppendieck et al. 2007a, Poppendieck et al. 2007b).
`o` **Carbonyl by-products** (Morrison et al. 2021; secondary sources cited Poppendieck et al. 2007b).
`o` **C1 to C10 saturated aldehydes** (e.g., formaldehyde, acetaldehyde), acetone, and organic acids (Namdari et al. 2021) is widely documented.
`o` In addition to aldehydes, ozone may also increase indoor concentrations of formic acid, both of which can irritate the lungs if produced in sufficient amounts (US EPA 2021).
`o` Additionally, by-product formation has been observed after exposure to significantly **reduced ozone concentrations (0.1 ppm)** (Morrison et al. 2021; secondary sources cited Morrison and Nazaroff 2002; Morrison et al. 1998).

“Due to its toxicological properties and its capability to degrade several materials, the optimal use of ozone for the disinfection of air and surfaces is in the absence of humans, using a dose and time of usage sufficient to destroy viruses, but having minimal degradation effects on materials”. (Grignani et al. 2021. p.29.)

### 3.3 Areas of uncertainty

***Q1: What is the evidence for the surface survival of coronaviruses?***

- There is a **lack of review evidence** retrieved that specifically reported on the survival **of** SARS-CoV-2 **in** real world settings.
- **Laboratory-based virus** survival experiments use **different virus titres** that contained SARS-CoV-2 in much **higher ratios** than in a **real-world setting** (NCCMT 2021), therefore, findings should be interpreted with caution.
- Further study is needed on whether contact with contaminated surfaces had any importance in SARS-CoV-2 infections (NHLKS 2020).

***Q2a: What is the evidence for the potential efficacy of ozone as an air or surface disinfectant against coronaviruses?***

- There are currently **no ozone disinfection investigations** directly examining **SARS-CoV-2** (Morrison et al. 2020).
- Research has all been conducted in **laboratory-based settings** or after samples have been **purposely placed on surfaces in situ** (hotel rooms, cruise liner cabin, offices or healthcare environments).

***Q3a: What is the effectiveness of no-touch automated ozone disinfection methods for decontamination against SARS-CoV-2?***

- There were no reviews or primary studies that investigated the effectiveness of ozone machines against SARS-CoV-2 in any real-world setting.

***Q4a: What are the potential health effects of ozone for children and young people?***

- There is no direct evidence for the effects on health directly related to the use of ozone machines in educational settings for children, young people and staff.

***Q4b: What are the benefits and harms of using ozone machines?***

- There is no direct evidence for the harms and benefits directly related to the use of ozone machines in educational settings for children, young people and staff.

## 4. Options for further work

- A rapid evidence summary of gaseous chlorine dioxide for disinfection of SARS-Cov- 2.
- A rapid review of the efficacy and effectiveness of ozone machines as a method of decontamination in indoor settings as previous reviews have been narrative in nature. The one systematic review that has been conducted in this area was for the decontamination using ozone in a sealed chamber and the authors did not undertake quality appraisal.

## 6. Methods used in this Rapid Evidence Summary

COVID-19 specific and general repositories of evidence reviews; the reference databases PubMed, Medline, Embase and Web of Science; and websites of key originations were searched on 8^th^ to 10^th^ September 2021. An audit trail of the search process (and search terms used) is provided within the resource list (Appendix). Searches were limited to English-language publications and did not include searches for primary studies if secondary research relevant to the question was found.

Search hits were screened for relevance by a single reviewer. Priority was given to robust evidence synthesis using minimum standards (systematic search, study selection, quality assessment, appropriate synthesis). The secondary research identified was not retrieved as full text or formally quality assessed. The included research may vary considerably in quality and the degree of such variation could be investigated during rapid review work which may follow-on. Citation, recency, evidence type, document status and key indications were tabulated for all relevant secondary research identified in this process.

As secondary evidence was limited, a further targeted search for primary studies was conducted to inform options for further work. Findings from such studies have not been tabulated but an indication is given of the amount of literature for different aspects of the question.

|  |  |
| --- | --- |
| **Date of Search** | September 2021 |
| **Search Concepts Used** | Ozone OR ultraviolet OR hydrogen peroxide OR atomat* OR no touch AND surface OR AIR AND clean* OR disinfect OR decontam* or inactiv* |
| **Search Completed by** | Elizabeth Gillen, Deborah Edwards, WCEBC |

## Data Availability

No primary data is associated with this work. All research studies included are available online in relevant databases, such as Medline, Web of Sciences, etc.

https://healthandcareresearchwales.org/about-research-community/wales-covid-19-evidence-centre

## 7. Results

**Table 1.** Summary of review evidence identified

| Evidence type | Total identified | Comments |
| --- | --- | --- |
| Systematic reviews (SRs)<br>- Q1: Surface survival<br>- Q2a: Efficacy of ozone<br>- Q2b: Efficacy of Light-based technologies<br>- Q2c: Efficacy of hydrogen peroxide vapour<br>- Q3a: Effectiveness of ozone<br>- Q3b: Effectiveness of Light-based technologies<br>- Q3c: Effectiveness of hydrogen peroxide vapour<br>- Q4a: Heath<br>- Q4b: Harms | 3<br>1<br>3<br>1<br>0<br>1<br>0<br>0<br>0 |  |
| Rapid reviews (RRs)<br>- Q1: Surface survival<br>- Q2a: Efficacy of ozone<br>- Q2b: Efficacy of Light-based technologies<br>- Q2c: Efficacy of hydrogen peroxide vapour<br>- Q3a: Effectiveness of ozone<br>- Q3b: Effectiveness of Light-based technologies<br>- Q3c: Effectiveness of hydrogen peroxide vapour<br>- Q4a: Heath<br>- Q4b: Harms | 2<br>0<br>0<br>1<br>1<br>1<br>1<br>0<br>0 |  |
| Narrative review (NRs)<br>- Q1: Surface survival<br>- Q2a: Efficacy of ozone<br>- Q2b: Efficacy of Light-based technologies<br>- Q2c: Efficacy of hydrogen peroxide vapour<br>- Q3a: Effectiveness of ozone<br>- Q3b: Effectiveness of Light-based technologies<br>- Q3c: Effectiveness of hydrogen peroxide vapour<br>- Q4a: Heath<br>- Q4b: Harms | 0<br>6<br>4<br>0<br>1<br>1<br>1<br>1<br>7 |  |
| Evidence Synthesis<br>Q4a: Heath | 1 |  |
| Protocols for reviews that are underway<br>- Q1: Surface survival<br>- Q2a: Efficacy of ozone<br>- Q2b: Efficacy of Light-based technologies<br>- Q2c: Efficacy of hydrogen peroxide vapour<br>- Q3a: Effectiveness of ozone<br>- Q3b: Effectiveness of Light-based technologies<br>- Q3c: Effectiveness of hydrogen peroxide vapour<br>- Q4a: Heath<br>- Q4b: Harms | 0<br>0<br>1<br>1<br>0<br>2<br>0<br>1<br>0 |  |
| Science Brief<br>- Q1- Surface survival | 1 |  |
| Primary Studies<br>- Q2a Efficacy of ozone<br>- Q3a: Effectiveness of ozone<br>- Q4b: Harms | 6<br>1<br>5 |  |
| Secondary sources (citations within reviews) |  |  |
| - Q1: Surface survival | 0 |  |
| - Q2a: Efficacy of ozone | 0 |  |
| - Q2b: Efficacy of light-based technologies | 0 |  |
| - Q2c: Efficacy of hydrogen peroxide vapour | 1 | letter |
| - Q3a: Effectiveness of ozone | 0 |  |
| - Q3b: Effectiveness of Light-based technologies | 0 |  |
| - Q3c: Effectiveness of hydrogen peroxide vapour | 0 |  |
| - Q4a: Heath | 0 |  |
| - Q4b: Harms | 8 | Not tabled |
| Tertiary sources |  |  |
| - Q3b: Efficacy of light-based technologies | 3 | 2 SAGE/1 TAG |
| - Q3c: Efficacy of hydrogen peroxide vapour | 3 | 2 SAGE/1 TAG |
| - Q4a: Heath | 2 | SAGE / US EPA |
| - Q4b: Harms | 1 | US EPA |

A more detailed summary of included evidence can be found in Tables 2 - 10

**Table 2:** Summary of secondary research for surface survival of coronaviruses

| Resource | Citation | Recency<br>(Search dates) | Evidence<br>Type* | Status | Key findings from abstracts | Reviewer comments |
| --- | --- | --- | --- | --- | --- | --- |
| VA-ESP | National Collaborating<br>Centre for Methods and<br>Tools (2021)<br>Rapid Review Update 1:<br>What is known about how<br>long the virus can survive<br>with potential for infection<br>on surfaces found in<br>community settings? March<br>5<sup>th</sup> 2021 | 31 July 2020 to<br>31 Dec 2020 | RR | Published | <u>Focus</u><br>What is known about how long the virus can survive with potential for infection on surfaces found in community settings?<br><u>Setting</u><br>Inanimate surfaces commonly found in community settings (clinical and hospital settings excluded)<br><u>Key findings</u><br>**There is consistent evidence that fragments of SARS-CoV-2 can be detected on surfaces in community settings for up to seven days** (certainty of evidence is **moderate (GRADE)**). However, most of these studies did not distinguish between live virus and dead virus or viral fragments. Only one study measured viable virus (that which has potential to infect) in samples and found none to be present<br>**While viral fragments can be detected on surfaces, these fragments may not be viable** (certainty of evidence is **low (GRADE)**).<br>**Cleaning procedures consistently decreased or eliminated detection of SARS-CoV-2 fragments** (certainty of the evidence is **moderate (GRADE)**).<br>**Laboratory-based studies indicate SARS-CoV-2 can remain viable longer on smoother surfaces such as plastic or steel than cardboard or cotton.** | <u>Number of studies</u><br>Completed syntheses (n=7)<br>In progress syntheses (n=2)<br>Single studies (n=32)<br><u>Quality appraisal</u><br>AMSTAR-1<br>JBI Checklist for Prevalence Studies<br>GRADE<br><u>Key references</u><br>*Completed syntheses:*<br>Bedrosian et al. 2020<br>Bueckert et al. 2020<br>Fernández-Raga et al. 2020<br>Meyerowitz et al. 2020<br>Usher Institute 2020<br>Fiorillo et al. 2020<br>National Academies of Sciences, Engineering, and Medicine 2020<br>*In progress:*<br>Dall Nora et al. 2020<br>Deliga Schroder et al. 2020 |
|  |  |  |  |  | However often with starting concentrations much higher than found in the environment. There is wide variation in the length of times reported but there is indication of increased stability at lower temperatures (such as 4°C) and more rapid decay with increasing temperatures ((evidence cannot be rated on GRADE)) |  |
| VA-ESP | Bedrosian et al. (2021)<br>A systematic review of surface contamination, stability, and disinfection data on SARS-CoV-2 (Through July 10, 2020).<br>Environ. Sci. Technol. 55: 4162-73 | 22 Jan 2020 to 10 July 2020 | SR | Published | <u>Focus</u><br>A systematic review of hygiene intervention effectiveness against SARS-CoV-2<br><u>Setting</u><br>Healthcare (patient room; non-COVID-patient room); Household; Non-household accommodation; Other shared (laboratory; outdoor)<br><u>Key findings</u><br>*Surfaces contamination:* in the community: 2.5% of household surfaces positive to SARS-CoV-2; 14% in non-household accommodation; and 14% in outdoor settings (including 25% high-touch surfaces and 23% hard furniture).<br>*Surface stability:* SARS-CoV-2 half-life: 2.3-17.9 hours on stainless steel; 2.3-15.3 hours on plastic; 2.3-15.3 hours on nitrile. Half-life decreases as temperature and humidity increase<br>*Surface disinfection:* A 99.9% virus reduction can be obtained with sunlight, ultraviolet light, ethanol, hydrogen peroxide and hypochlorite. | <u>Number of studies</u><br>Surface contamination (n=35)<br>Surface stability (n=16)<br>Surface disinfection (n=27)<br><u>Quality appraisal</u><br>Did not systematically assess risk of bias on the basis of using preprints.<br><u>General comments</u><br>Summarised as part of the rapid review conducted by NCCMT 2021 |
| VA-ESP<br>L*OVE | Marzoli et al. (2021)<br>A systematic review of human coronaviruses | Up to 6 Nov 2020 | SR | Published | <u>Focus</u> | <u>Number of studies</u><br>Total of full text reviewed (n=18) |
|  | survival on environmental surfaces. Sci Total Environ. 15(778): 146191. |  |  |  | To summarize all the evidence on surface survival of coronaviruses infecting humans<br><u>Setting</u><br>Experimental setting<br><u>Key findings</u><br>The longest coronavirus survival time is 28 days at room temperature on different surfaces: polymer banknotes, vinyl, steel, glass, and paper banknotes. **Concerning SARS-CoV-2 human infection from contaminated surfaces, dangerous viral load on surfaces for up to 21 days was determined on polymer banknotes, steel, glass and paper banknotes**<br>For viruses other than SARS-CoV-2, the longest period of survival was 14 days, recorded on glass. Environmental conditions can affect virus survival, and indeed, low temperatures and low humidity support prolonged survival of viruses on contaminated surfaces independently of surface type. Furthermore, it has been shown that exposure to sunlight significantly reduces the risk of surface transmission | <u>Quality appraisal</u><br>Conducted but no tool mentioned<br><u>General comments</u><br>Although studies are increasingly investigating the topic of coronavirus survival, it is difficult to compare them, given the methodology differences. For this reason, it is advisable to define a reference working protocol for virus survival trials, but, as an immediate measure, there is also a need for further investigations of coronavirus survival on surfaces |
| Organisational websites | National Health Library and Knowledge Service (2020) How long can the COVID-19 virus exist on surfaces? What role do contaminated surfaces play in the chain of transmission? What infection control precautions should be implemented? HSE | Up to 6 July 2020 | RR | Published | <u>How long can the COVID-19 virus exist on surfaces?</u><br>Human coronaviruses such as Severe Acute Respiratory Syndrome (SARS) coronavirus, Middle East Respiratory Syndrome (MERS) coronavirus or endemic human coronaviruses (HCoV) can persist on inanimate surfaces such as metal, glass or plastic for up to 9 days | <u>Number of studies</u><br>40 papers found in reference list, including international guidance<br><u>Quality appraisal</u><br>None<br><u>Key references:</u><br>Kampf et al. 2020a |
|  | Summary of Evidence:<br>COVID-19. 6 July 2020 |  |  |  | SARS-CoV-2 was more stable on plastic and stainless steel than on copper and cardboard, and that viable virus was detected up to 72 hours after application to these surfaces although the virus titre was greatly reduced<br>The persistence time on inanimate surfaces varied from minutes to up to one month, depending on environmental conditions, and that SARS-CoV-2 can be sustained in air in closed unventilated buses for at least 30 minutes without losing infectivity<br><u>What role do contaminated surfaces play in the chain of transmission?</u><br>Virus particles in the air and on fomites are exposed to a range of environmental conditions that influence their persistence. Relative humidity, fomite material and air temperature can greatly impact enveloped virus inactivation rates. The importance of indirect contact transmission involving contamination of inanimate surfaces is uncertain and warrants further study<br><u>What infection control precautions should be implemented?</u><br>The CDC recommend cleaning and disinfection of high-touch surfaces daily in household common areas: e.g. tables, hard-backed chairs, doorknobs, light switches, remotes, handles, desks, toilets, sinks<br>Exposure to natural sunlight, the use of antimicrobial copper surfaces, the application of a modified antimicrobial coating on surfaces and sensor taps and no door handles may be effective supplements to standard hygiene practices and present additional<br>opportunities for controlling the transmission of COVID-19 from contaminated fomites | Kampf et al. 2020b<br>Van Doremalen et al. 2020<br>Ren et al. 2020<br>Wiggington and Boehm 2020<br>Ratnesar-Shumate et al. 2020<br>ECRI 2020<br>Ikner et al. 2020<br>CDC 2020 [Updated 15 June 2021]<br>CDC 2020 [Updated 17 June 2021] |
| Organisational websites | Centre for Disease Control and Prevention (2021) Science Brief: SARS-CoV-2 and surface (fomite) transmission for indoor community environments. CDC, April 2021 | Not reported | Science Brief | Published | <u>Risk of infection</u><br>The principal mode by which people are infected with SARS-CoV-2 (the virus that causes COVID-19) is through exposure to respiratory droplets carrying infectious virus. It is possible for people to be infected through contact with contaminated surfaces or objects (fomites), but the risk is generally considered to be low<br><u>Surface survival</u><br>When accounting for both surface survival data and real-world transmission factors, the risk of fomite transmission after a person with COVID-19 has been in an indoor space is minor after 3 days (72 hours), regardless of when it was last cleaned. Researchers have found that 99% reduction in infectious SARS-CoV-2 on non-porous surfaces can occur within 3 days<br><u>Effectiveness of cleaning and disinfection</u><br>Disinfection is recommended in indoor community settings where there has been a suspected or confirmed case of COVID-19 within the last 24 hours. The risk of fomite transmission can be reduced by wearing masks consistently and correctly, practicing hand hygiene, cleaning, and taking other measures to maintain healthy facilities | <u>Key References</u><br>Harvey et al 2021. Longitudinal monitoring of SARS-CoV-2 RNA on high-touch surfaces in a community setting. Environ Sci Technol Lett. 8(2):168-75.<br>Pitol and Julian 2021. Community transmission of SARS-CoV-2 by fomites: Risks and risk reduction strategies. Environ Sci Technol Lett. Jan 6 : acs.estlett.0c00966.<br>Biryukov et al 2020. Increasing temperature and relative humidity accelerates inactivation of SARS-CoV-2 on surfaces. mSphere, 5(4): e00441-20, 2020.<br>Chin et al 2020. Stability of SARS-CoV-2 in different environmental conditions. Lancet Microbe. 1(1): e10, 2020.<br>Kratzel et al. 2020. Temperature-dependent surface stability of SARS-CoV-2. J Infect. 81(3): 452-82<br>Liu et al. 2021. Stability of SARS-CoV-2 on environmental surfaces and in human excreta. J Hosp Infect. Jan (107):105-7. |
| L*OVE<br>PubMed | Kampf et al. (2020a)<br>Persistence of coronaviruses on inanimate surfaces and their inactivation with biocidal agents. J Hosp Infect. 104(3): 246–251.<br>Kampf (2020)<br>Potential role of inanimate surfaces for the spread of coronaviruses and their inactivation with disinfectant agents. Infect Prev Pract. 2(2): 100044. | 28 January 2020 | SR | Published | <u>Focus</u><br>To review the literature on all available information about the persistence of human and veterinary coronaviruses on inanimate surfaces as well as inactivation strategies with biocidal agents used for chemical disinfection<br><u>Setting</u><br>Healthcare facilities, although it is unclear if included studies looked at care setting or were experimental<br><u>Methods of disinfection</u><br>Biocidal agents and chemical disinfection*<br><u>Key findings</u><br>Severe Acute Respiratory Syndrome coronavirus, Middle East Respiratory Syndrome coronavirus or endemic human coronaviruses can persist on inanimate surfaces like metal, glass or plastic for up to 9 days | <u>Number of studies</u><br>Total of full text reviewed (n=22)<br><u>Quality appraisal</u><br>None |

Key: \* RR Rapid review; SR systematic review; NR: narrative review

**Table 3:** Summary of secondary research for the potential efficacy of ozone as an air or surface disinfectant against coronaviruses

| Resource | Citation | Recency (Search dates) | Evidence Type* | Status** | Key findings from abstracts | Reviewer comments |
| --- | --- | --- | --- | --- | --- | --- |
| WHO | Grignani et al (2021)<br>Safe and effective use of ozone as air and surface disinfectant in the conjuncture of Covid-19.<br>Gases. 1(1)<br>10.3390/gases1010002 | Between June and Nov 2020 | NR | Published | <u>Focus</u><br>To conduct a literature review on the ozone virucidal efficacy in order to individuate the optimal conditions (concentration, contact time, microclimate) for its possible use as disinfectant for indoor environments and Personal Protective Equipment (PPE), with particular reference to SARS-CoV-2<br><u>Settings</u><br>Indoor environments<br><u>Viruses and Bacterium</u><br>Herpes, poliovirus, norovirus, influenza, influenza A, respiratory syncytial virus, ssDNA, ssRNA, dsDNA, Enveloped dsRNA, phages, different virus not specified, SARS-CoV-2<br><u>Key Findings</u><br>Ozone is a powerful oxidant reacting with organic molecules, and therefore has bactericidal, virucidal, and fungicidal actions<br>Ozone can be generated in situ by means of small, compact ozone generators, using dried ambient air as a precursor. It should be injected into the room that is to be disinfected until the desired ozone concentration is reached; after the time needed for the disinfection, its | <u>Number of studies</u><br>Ozone virucidal efficacy (n=18)<br>Disposable masks and PPE for reuse (n=20)<br><u>Quality appraisal</u><br>None<br><u>Safety and residuals</u><br>Limited data are available, but they indicate significant damage to rubber products and surface coatings, while textiles and other polymeric materials seem not to be affected in the range of atmospheric concentrations (Lee et al 1996)<br>Ozone is also able to fade dyes on nylon and acetate, and most of the natural dyes and dye-based pigments used by artists. It also plays a role in the process of the corrosion of metals like copper and aluminium (Druzik 1985)<br>Due to its toxicological properties and its capability to degrade several materials, the optimal use of ozone for the disinfection of air and surfaces is in the absence of humans, |
|  |  |  |  |  | concentrations must be reduced to the levels required for the workers' safety | using a dose and time of usage sufficient to destroy viruses, but having minimal degradation effects on materials. |
| Web of Science | Bayarri et al. (2021)<br>Can ozone inactivate SARS-CoV-2? A review of mechanisms and performance on viruses.<br>Journal of Hazardous Materials. 415(2021): 125658 | Not reported<br>*papers included from 1982 to 2020 | SR | Published | <u>Focus</u><br>To carry out a critical review of all the published works related to O3 gas-phase applications in order to discern the effectiveness of this method as a virucidal agent<br><u>Setting</u><br>Experimental setting (sealed chambers)<br><u>Viruses</u><br>28 different viruses of the 29 tested, including SARS-CoV-2 surrogates<br><u>Methods of disinfection</u><br>Ozone in gas-phase<br><u>Key Findings</u><br>It could also be concluded that **gaseous ozone** can be indeed an **effective disinfectant**, successfully **inactivating viruses such as influenza A H1N1, MERS-CoV, SARS-CoV-1 or even SARS-CoV-2 in aerosols or fomites**. In reviewed works, low ozone exposures, just around 0.1–0.4 mg L<sup>-1</sup> min, achieve about 4 log<sub>10</sub> of inactivation in aerosols, while exposures between 1 and 4 mg L<sup>-1</sup> min may be needed to guarantee an inactivation of 3–4 log<sub>10</sub> in different fomites | <u>Number of studies</u><br>Virus inactivation in aerosols (n=6)<br>Disinfection on surfaces or fomites (n=14)<br><u>Quality appraisal</u><br>None<br><u>General comments</u><br>The authors referred to it as a critical review. Although they mentioned databases searched, and that they screened over 200 papers (no exact number mentioned)<br><u>Safety</u><br>No mention of safety issues in real world environment |
| MEDLINE | Blanco et al. (2021)<br>Ozone potential to fight against SAR-COV-2 | Not reported | NR | Published | <u>Focus</u> | <u>Number of studies</u><br>Not stated |
|  | pandemic: facts and research needs.<br>Environmental Science and Pollution Research (2021) 28:16517–31 | *papers mentioned from 1977 to 2020 |  |  | To review the most relevant results of virus disinfection by the application of gaseous ozone<br>Disinfection treatments of both air and surfaces<br><u>Setting</u><br>Experimental setting<br>(from boxes to larger rooms)<br><u>Methods of disinfection</u><br>Ozone in gas-phase<br><u>Viruses</u><br>A variety of different viruses including SARS-CoV-2 surrogates<br><u>Key findings</u><br>In small chambers, 10–20 mg ozone/m<sup>3</sup> over 10 to 50 min can be sufficient to significantly reduce the virus load of personal protection equipment.<br>In large rooms, 30 to 50 mg ozone/m<sup>3</sup> would be required for treatments of 20–30 min. Maximum antiviral activity of ozone is achieved at high humidity, while the same ozone concentrations under low RH could result inefficient | <u>Quality appraisal</u><br>Critical analysis mentioned but unclear how this was conducted<br><u>General comments</u><br>No methods mentioned, and no number of papers. I put review down, as no explicit mention on this being a narrative review<br><u>Advantages</u><br>As a gas, it can penetrate all areas within a room, including crevices, fixtures, fabrics, and the under surfaces of furniture, much more efficiently than liquid sprays, aerosols, or ultraviolet light.<br><u>Safety</u><br>The ability of ozone to corrode certain materials after prolonged exposure has not been observed in the few data available on PPE disinfection<br>Gaseous ozone is an extremely oxidizing agent, and severe effects can be observed when ozone is applied to devices made of natural rubber or derived composites |
| WHO<br>HERO | Morrison et al. (2021)<br>Critical review and research needs of ozone applications related to virus inactivation: | Not reported | NR | Published | <u>Focus</u><br>To identify the exposure requirements for virus inactivation and important safety considerations for applications within the | <u>Number of studies</u><br>Not stated<br><u>Quality appraisal</u> |
|  | potential implications for SARS-CoV-2. Ozone: Science & Engineering. 43(1): 2-20 |  |  |  | built environment (i.e. occupied/unoccupied spaces, air/water/wastewater treatment) and healthcare settings (i.e. ozone therapy, dentistry, handwashing, treatment of personal protection equipment.<br><u>Setting</u><br>Experimental and healthcare; water treatment; air cleaning in occupied and unoccupied spaces<br><u>Methods of disinfection</u><br>Ozone (in gas and aqueous form)<br><u>Viruses</u><br>A variety of different viruses including SARS-CoV-2 surrogates<br><u>Key findings</u><br>Ozone disinfection has demonstrated high efficacy against enveloped and non-enveloped viruses, including viruses similar in morphology to SARS-CoV-2.<br>With proper guidance, there are many potential applications for the use of ozone within the built environment, beyond the ongoing pandemic. However, there are currently many needs which must be addressed to ensure safe and effective use of ozone in gaseous and aqueous forms | None<br><u>General comments</u><br>No methods section, no explicit statement on search and screening processes and papers found<br><u>Safety and residuals</u><br>One potential concern is the accelerated decay of indoor materials exposed to the ozone concentrations required for disinfection. For instance, cracking of rubber has been observed within 24 hours of exposure to ozone at 0.5 ppm (Kamaruddin and Muhr 2018)<br>Additionally, ozone may react with a range of materials commonly anticipated in the indoor environment including paint, linoleum, carpet, paper, wood and semi-volatile organics adsorbed to surfaces (Weschler 2000)<br>Following repeated exposure to ozone, building materials may lose colour, and physically degrade.<br><u>Formation of by-products</u><br>As a result of heterogeneous (gas/solid) reactions, a variety of organic by-products may be formed after exposure to ozone (Morrison and Nazaroff 2002; Morrison et al. 1998; |
|  |  |  |  |  |  | Poppendieck et al. 2007a, Poppendieck et al. 2007b).<br>Carbonyl by-products are commonly reported to be present across a range of different surface types. Paper, office partition materials, and medium density fibreboard (e.g. furniture material) exhibited the highest overall by-product formation of the studied materials. By-products (e.g. nonanal) were shown to be persistent for months following exposure to ozone at emission rates which exceed odour detection concentrations (Poppendieck et al. 2007b).<br>Additionally, by-product formation has been observed after exposure to significantly reduced ozone concentrations (0.1 ppm) (Morrison and Nazaroff 2002; Morrison et al. 1998). |
| HERO | Alimohammadi and Naderia (2021)<br>Effectiveness of ozone gas on airborne virus inactivation in enclosed spaces: A review study. Ozone: Science & Engineering. 43(1): 21-31 | Not reported | NR | Published | <u>Focus</u><br>To bring attention to the ozonizing of indoor air as a novel treatment for the inactivation of viruses<br><u>Setting</u><br>Experimental but not explicitly mentioned<br><u>Methods of disinfection</u><br>Ozone gas<br>Viruses | <u>Number of studies</u><br>Not stated<br><u>Quality appraisal</u><br>None<br><u>General comments</u><br>No methods section, no explicit statement on searches, screening and study selection<br>Safety |
|  |  |  |  |  | Enveloped viruses (e.g. SARS-CoV-2)<br><u>Key findings</u><br>Enveloped viruses (e.g. SARS-CoV-2) are more sensitive to oxidizing agents such as ozone than to non-enveloped viruses. Furthermore, some viruses such as coronaviruses have cysteine containing a sulfhydryl group that reacts with ozone gas<br>The study indicated that more free radicals will be formed when air humidity is higher, which could lead to higher virus inactivation<br>Air disinfection by ozone gas can be a promising approach for the viral deactivation of contaminated spaces in hospitals, healthcare centres, dental offices, sport clubs, hotels and transportation sector, as well as all other places where viral disease outbreaks occur | The only significant drawback is its ability to corrode certain materials, such as natural rubber, long-term exposure, and its potential toxicity to humans<br>By ensuring that the room is temporarily closed to people during treatment, health risks are eliminated and sealed to prevent gas from escaping into the environment. If necessary, the sensitive material can be temporarily covered or removed |
| PubMed | Otter et al. 2013<br>The role of 'no-touch' automated room disinfection systems in infection prevention and control. J Hosp Infect 83(1):1e13. | Not reported | NR | Published | <u>Focus</u><br>Explores microbiological efficacy of automated room disinfection systems<br><u>Setting</u><br>In vitro and In situ<br><u>Methods of disinfection</u><br>Aerosolized hydrogen peroxide<br>H<sub>2</sub>O<sub>2</sub> vapour / Hydrogen Peroxide vapour<br>Ultraviolet C radiation<br>Pulsed-xenon UV<br>Gaseous ozone<br>Chlorine dioxide<br>Fogging<br><u>Viruses</u> | <u>Number of included studies</u><br>In Vitro<br>- Aerosolized hydrogen peroxide (n=12)<br>- H<sub>2</sub>O<sub>2</sub> vapour (n=17)<br>- UVC (n=8)<br>- Pulsed-xenon (n=2)<br>In Situ<br>- Hydrogen Peroxide vapour (n=14)<br>- Aerosolized hydrogen peroxide (n=5)<br>- UVC (n=4)<br>- Pulsed-xenon (n=2) |
|  |  |  |  |  | Not specified<br><u>Key Findings</u><br>**Ozone is efficacious for room decontamination.** Substantially lower reductions were achieved at lower relative humidity. The requirement for high humidity is a major practical limitation for ozone-based systems<br>**Chlorine dioxide** has a high level of efficacy against a range of pathogens. However, concerns about safety and material compatibility mean that it is unlikely to be used in healthcare settings | <u>Safety during operation</u><br>Ozone is toxic to humans, with a safe exposure level in the United Kingdom and United States of <0.1 ppm (compared with 1 ppm for hydrogen peroxide), so effective containment of the gas, monitoring for leakage, and measurements to assure that the room is safe to enter are necessary for these systems in the healthcare setting<br><u>Safety and residuals</u><br>Data on the compatibility of this process with hospital materials are needed, given ozone's known corrosive properties (Davies et al 2011)<br><u>Key References</u><br>Sharma and Hudson 2008<br>Moat et al. 2009.<br>Zoutman et al. 2011 |
| Back chaining | Davies et al 2011.<br>Gaseous and air decontamination technologies for Clostridium difficile in the healthcare environment. J Hosp Infect. 77:199–203. | Not reported | NR | Published | <u>Focus</u><br>A review of gaseous and air decontamination technologies for Clostridium difficile in the healthcare environment<br><u>Setting</u><br>Healthcare environment<br><u>Viruses</u><br>Clostridium difficile<br><u>Methods of disinfection</u> | <u>Number of studies</u><br>Not reported<br><u>Quality appraisal</u><br>None<br><u>Safety after operation</u><br>An exposure limit over 15 min has been set at 0.2 ppm at which concentration some people can still experience respiratory symptoms, but at which concentration, ozone |
|  |  |  |  |  | Hydrogen peroxide, chlorine dioxide, ozone, Ub based technologies<br><u>Key Findings</u><br>**Gaseous hydrogen peroxide may be a useful** additional tool in the attempts to reduce environmental contamination, but further studies are still needed to determine its practical use in reducing transmission in the hospital setting | has limited microbicidal efficacy<br><u>Safety and residuals</u><br>However, since ozone is toxic and a potent oxidiser that corrodes metals, it has not been widely investigated in the hospital environment<br>At humidifies of >80%, ozone will attack and degrade rubber and therefore compatibility with local materials should be considered (A. Bennett, personal communication). |

Key: \* NR: Narrative review; RR Rapid review; SR systematic review

**Table 4:** Summary of primary studies for the potential efficacy of ozone machines as an air or surface disinfectant against coronaviruses

| Resource | Citation | Focus | Key findings from abstracts |
| --- | --- | --- | --- |
| **Laboratory based or purposely placed on surfaces in situ** |  |  |  |
| Back chaining | Hudson et al. (2007)<br>Inactivation of norovirus by ozone gas in conditions relevant to healthcare. J Hosp Infect. 66:40e5. | <u>Aim</u><br>To evaluate the ability of ozone gas to inactivate Norovirus and its animal surrogate feline calicivirus in dried samples placed at various locations within a hotel room, a cruise liner cabin and an office<br><u>Viruses</u><br>Norovirus and its animal surrogate feline calicivirus<br><u>Setting</u><br>In dried samples placed at various locations within a hotel room, a cruise liner cabin and an office | <u>Effectiveness</u><br>Our results show that Norovirus can be inactivated by exposure to ozone gas from a portable commercial generator in settings such as hotel rooms, cruise ship cabins and healthcare facilities<br><u>Advantages</u><br>Ozone gas has several advantages as a practical antiviral agent. It can effectively penetrate every part of a room, including sites that might prove difficult to gain access to with conventional liquids and manual cleaning procedures. For example, in our tests, virus deposited under the table or adsorbed to fabric taped to a window were just as vulnerable to attack as virus placed in more accessible sites<br>he gas is easy and economical to produce, and is a natural compound which decays quickly back to oxygen with a half-life of about 20 min. The use of a catalytic converter (scrubber) considerably speeds up the removal of the gas. In addition, in the event of possible malfunction during application, the gas is readily detected by smell and hence can be avoided<br><u>Safety during operation</u><br>Its major disadvantage is its potential toxicity at high concentration, which precludes its use in areas continuously populated by people. In practice this means it can only be used in rooms that can be sealed off or quarantined for the duration of the treatment. Since the standard protocol requires less than an hour to perform, however, this should not be a barrier to utilization given its potential efficacy |
| PubMed | Franke et al. (2021)<br>An automated room disinfection system using ozone is highly active against | <u>Aim</u><br>The determine the effectiveness of a fully automatic room decontamination system based on ozone | <u>Effectiveness</u><br>The ozone-based room decontamination device achieved virucidal efficacy (reduction factor >4 log10) against both |
| | surrogates for SARS-CoV-2.<br>J Hosp Infect.12:108-113. | <u>Viruses</u><br>Bacteriophage $\Phi 6$ (phi 6) and bovine coronavirus L9, as surrogate viruses for the pandemic coronavirus SARS-CoV-2<br><u>Surfaces</u><br>**Various surfaces (ceramic tile, stainless steel surface and furniture board)** were soiled with the surrogate viruses and placed at two different levels in a gas-tight test room | surrogate organisms regardless of the different surfaces and positions confirming a high activity under the used conditions<br>In summary, we found that **ozone in combination with high humidity as generated by an automated room decontamination system has a high activity against SARS-CoV-2 surrogate viruses bacteriophage F6 and BCoV on different solid surfaces in the hospital environment**, confirming the process as a virucidal disinfection<br><u>Safety during operation</u><br>However, due to toxicity of ozone, doors and ventilation diffusers must be strictly sealed to prevent unintentional dissemination resulting in an additional workload for the operating person<br>Additionally, due to the generated water aerosol, smoke detectors must also be covered to avoid unwanted alarms<br>During the disinfection cycle, a concept is needed to prevent unauthorized room entrance during the disinfection process |
| PubMed | Uppal et al. (2021)<br>Inactivation of human coronavirus by FATHHOME's dry sanitizer device: rapid and eco-friendly ozone-based disinfection of SARS-CoV-2. Pathogens. 14;10(3):339. | <u>Aim</u><br>To evaluate the virucidal efficacy of FATHHOME's self-contained, ozone-based dry-sanitizing device, by dose and time response assessment<br><u>Viruses</u><br>We tested inactivation of human coronavirus, HCoV-OC43, a close genetic model of SARS-CoV-2<br><u>Surfaces</u><br>On porous (N95 filtering facepiece respirator/FFR) and nonporous (glass) surfaces | <u>Effectiveness</u><br>We started our assays with 20 ppm-10 min ozone exposure, and effectively reduced 99.8% and 99.9% of virus from glass and N95 FFR surfaces, respectively. Importantly, the virus was completely inactivated, below the detection limit (over 6-log<sub>10</sub> reduction) with 25 ppm-15 min ozone exposure on both tested surfaces<br>**As expected, a higher ozone exposure (50 ppm-10 min) resulted in faster inactivation** of HCoV-OC43 with 100% inactivation from both the surfaces.<br><u>Safety after operation</u><br>No residual ozone present after completion of the 5-min post exposure recapture cycle and no measurable increase in ambient ozone levels |
| PubMed | Steinmann et al. (2021)<br>Virucidal efficacy of an ozone-generating system for | <u>Aim</u> | <u>Effectiveness</u> |
|  | automated room disinfection.<br>J Hosp Infect. 16: 16-20 | The whole-room disinfectant device Sterisafe Pro, which creates ozone as a biocidal agent, was tested for its virucidal efficacy<br><u>Viruses</u><br>Inactivation of human adenovirus type 5 (AdV) and murine norovirus (MNV) (mandatory test viruses), modified vaccinia virus Ankara (MVA) and simian virus 40 (SV40)<br><u>Surfaces</u><br>Floor of a test room | All test virus titres were reduced after 150 and 300 min of decontamination, with mean reduction factors ranging from 2.63 (murine norovirus) to 3.94 (simian virus 40)<br>The enveloped virus (MVA) was more resistant to ozone than the non-enveloped viruses<br><u>Safety after operation</u><br>The risk of toxicity from aerosol exposure to ozone is well known, and a carcinogenic effect in animal models has been discussed. However, the new disinfection technology can be used in the hospital setting, as the indoor air concentration of ozone is lower and rooms are safe to enter after a completed cycle |
| PubMed | Knobling et al. (2021)<br>Evaluation of the effectiveness of two automated room decontamination devices under real-life conditions.<br>Front Public Health. 9:618263. | <u>Aim</u><br>To evaluate the effectiveness of the disinfection performance of a recently developed, fully automated system for generating ozone from atmospheric oxygen in combination with an integrated nebulizer for controlled increase of room humidity, was compared with a commercial nebulizer for generation of aerosolized hydrogen peroxide (aHP) under realistic conditions<br><u>Bacterium</u><br>Suspension of E. faecium ATCC 6057 with $5.0 \times 10^7$-$1.2 \times 10^8$ colony forming units (cfu)/mL was produced<br><u>Surface</u><br>Twenty-two contaminated surfaces were positioned in different areas in a fully furnished patient room with adjacent bathroom and anteroom High contaminated and low contaminated surfaces | <u>Effectiveness</u><br>Following the manufactures instructions, the ozone-based device displayed a bactericidal effect ($\log_{10} > 5$), whereas the aHP system failed for a high bacterial burden and achieves only a complete elimination of a realistic bioburden ($\log_{10} 2$). After increasing the exposure time to 30min, the aHP device also reached a bactericidal effect<br><u>Safety during operation</u><br>Both systems require a process time of more than 2h and also require time-consuming preparation (e.g., sealing doors, air diffusers, and smoke detectors with adhesive tape) and therefore cannot be used at all times<br><u>Safety and residuals</u><br>Ozone is a highly reactive and corrosive gas and in the future further investigations on material compatibility have to take place<br><u>Safety after operation</u><br>At the end of the process the active substance is completely degraded and the concentration of ozone prevailing in the room is continuously displayed on a mobile tablet computer and recorded in a standardized manner |
| PubMed | Dubuis et al. (2020)<br>Ozone efficacy for the control of airborne viruses: | <u>Aim</u> | <u>Effectiveness</u> |
|  | bacteriophage and norovirus models. PLoS ONE. 15(4):e0231164. | To test the efficacy of an air treatment using ozone and relative humidity for the inactivation of airborne viruses<br><u>Viruses and Bacterium</u><br>Four phages (φX174, PR772, MS2 and φ6) and one eukaryotic virus (murine norovirus MNV-1)<br><u>Surface</u><br>Biosafety level II cabinet | These findings suggest that ozone used at a low concentration is a powerful disinfectant for airborne viruses when combined with a high RH<br><u>Benefits</u><br>Using lower ozone concentrations is less costly because a high capacity ozone generator is not required.<br>Ozone concentrations of below 0.1 ppm may be feasible to treat the air inside unoccupied hospital rooms<br><u>Safety during operation</u><br>Because this gas is harmful to humans at concentrations above this value, patients and staff should not be present during air treatment in case the concentration exceeds 0.1 ppm<br>Another element to consider before implementing an air treatment plan involving ozone inside naturally ventilated rooms is the evaluation of the pressure inside the rooms. Negative pressure would prevent ozone leakage through the doors, but the majority of hospital rooms do not have this feature. Therefore, testing must be conducted for possible ozone leakage when doors are closed in order to evaluate the treatment's feasibility<br>For better protection, ozone destructors can be used and operated in the hallway near the closed door of the hospital rooms and inside them when the treatment is completed<br><u>Safety and residuals / by products</u><br>Additional investigations would also be needed regarding the interaction of ozone with other compounds found in hospital rooms, some of them released from furnishings and cleaning products |

**Table 5:** Summary of secondary research for the potential efficacy of light based technologies as an air and surface disinfectant against coronaviruses

| Resource | Citation | Recency<br>(Search dates) | Evidence<br>Type* | Status** | Key findings from abstracts | Reviewer comments |
| --- | --- | --- | --- | --- | --- | --- |
| **Efficacy of UV light based technologies** |  |  |  |  |  |  |
| L*OVE | Chiappa et al. (2021)<br>The efficacy of ultraviolet light-emitting technology against coronaviruses: a systematic review<br>J Hosp Infect. 114: 63–78. | Up to<br>22 Nov 2020<br>*papers included<br>from 1972 to 2020 | SR | Published | <u>Focus</u><br>To pool the available evidence on the efficacy of UV technologies against coronaviruses<br><u>Setting</u><br>Experimental setting (laboratory)<br><u>Methods of disinfection</u><br>UV light emitting technologies (including UVA, UVB, and UVC)<br><u>Viruses</u><br>SARS-CoV-1 (n=4)<br>MERS-CoV (n=1),<br>seasonal human coronaviruses (n=3)<br>**SARS-CoV-2 (n=6)**<br>animal coronaviruses (n=4)<br><u>Key findings</u><br>Overall, despite wide heterogeneity within included studies, **complete inactivation of coronaviruses on surfaces or aerosolized, including SARS-CoV-2, was reported to take a maximum exposure time of 15 min and to need a maximum distance from the UV emitter of up to 1 m.** Advances in UV-based technologies in the field of sanitation and their proved high virucidal potential against SARS-CoV-2 support their use for IPC in hospital and community settings and their contribution towards ending the COVID-19 pandemic | <u>Number of studies</u><br>Total of full text reviewed (n=18)<br><u>Quality appraisal</u><br>None on the basis of a lack of shared reporting standards for in vitro studies.<br><u>Safety during operation</u><br>Exposure to UV lamps is associated with health risks as **conventional UV light sources are recognized as a health hazard for humans, being both carcinogenic and cataractogenic**, involved in damage to eyes and skin<br>**Human exposure to artificial UV light should be avoided, thus most devices cannot be operated in the presence of people**, but used only in empty rooms and with motion sensors |
| LitCOVID | Hessling et al. (2020).<br>Ultraviolet irradiation doses for coronavirus inactivation – review and analysis of coronavirus photoinactivation studies. GMS Hyg Infect Control. 15: Doc08. | Not reported<br>*papers included from 1966 to 2020 | NR | Published | <u>Focus</u><br>To investigate the radiation dose necessary to inactivate SARS-CoV-2, the existing coronavirus photoinactivation results of the last 60 years have been reviewed and analysed<br><u>Setting</u><br>Experimental setting (tested on aerosol, droplets, surface, liquid)<br><u>Methods of disinfection</u><br>Ultraviolet irradiation (Almost all experiments were performed with mercury vapour lamps, with a peak emission at 254 nm (UVC). Individual investigations were performed with peak wavelengths at 222 nm (UVC), or 365 nm (UVA), or even with daylight)<br><u>Viruses</u><br>CoV, SARS-CoV, MERS-CoV<br><u>Key findings</u><br>The available data reveals large variations, which are apparently not caused by the coronaviruses but by the experimental conditions selected. If these are excluded as far as possible, it appears that coronaviruses are very UV sensitive. The upper limit determined for the log-reduction dose (90% reduction) is approximately 10.6 mJ/cm<sup>2</sup> (median), while the true value is probably only 3.7 mJ/cm<sup>2</sup> (median).<br>Since coronaviruses do not differ structurally to any great extent, the SARS-CoV-2 virus – as well as possible future mutations – will very likely be highly UV sensitive, so that **common UV**<br>**disinfection procedures will inactivate the new SARS-CoV-2 virus without any further modification** | <u>Number of studies</u><br>Not stated<br><u>Quality appraisal</u><br>None<br><u>General comments</u><br>Only PubMed and Google Scholar searched.<br>No search results mentioned and no details on screening processes<br><u>Safety</u><br>Not mentioned |
| LitCovid | Hadi et al. (2020)<br>Control measures for SARS-CoV-2: A review on light-based inactivation of single-stranded RNA viruses.<br>Pathogens. 9(9): 737. | Not reported | NR | Published | <u>Focus</u><br>To summarize the literature on light-based sanitization methods for the inactivation of ssRNA viruses in different matrixes (air, liquid, and solid).<br><u>Setting</u><br>Experimental setting (air, liquid and solid surfaces)<br><u>Methods of disinfection</u><br>Light-based (UV, blue, and red lights): mercury lamp (conventional UV), excimer lamp (UV), pulsed-light, and light-emitting diode (LED)<br><u>Viruses</u><br>Single-stranded RNA viruses (including Influenza A, human coronaviruses, feline calicivirus, Hepatitis A, Poliovirus-1, SARS-CoV)<br><u>Key findings</u><br>The rate of inactivation of ssRNA viruses in liquid was higher than in air, whereas **inactivation on solid surfaces varied with the type of surface**. The efficacy of light-based inactivation was reduced by the presence of absorptive materials.<br>**-Pulsed-light technologies** could inactivate viruses more quickly than conventional UV-C lamps.<br>-Large-scale use of **germicidal LED** is dependent on future improvements in their energy efficiency | <u>Number of studies</u><br>Not stated<br><u>Quality appraisal</u><br>None<br><u>General comments</u><br>No methods section; no explicit mention on review type; no mention of number of papers included, or screening processes<br><u>Safety</u><br>Ozone is generated, which limits the use of far-ultraviolet (FUV) due to its potential health risks.<br>Exposure to UVC range 253.7 nm has been associated with health risks, including **damage to the eyes and skin**.<br>**UV-C at 222 nm inflicted no damage on the eyes and skin of mice.** These data are still preliminary and further research is needed to ascertain the safety of UV light at 222 nm, especially its potential long-term effects on human health.<br>**Pulsed-light technologies** have improved safety (non-mercury). |
|  |  |  |  |  | **-Blue light** possesses virucidal potential in the presence of exogenous photosensitizers, although **femtosecond laser (ultrashort pulses)** can be used to circumvent the need for photosensitizers<br>**-Red light** can be combined with methylene blue for application in medical settings, especially for sanitization of blood products<br>Future modelling studies are required to establish clearer parameters for assessing susceptibility of viruses to light-based inactivation | **Blue light** is considered to be less harmful to mammalian cells than UV irradiation, albeit its photo-toxicity is wavelength-dependent. In addition to safety benefits, prolonged exposure of materials to blue light at 405 nm would induce less photo-degradation than UV irradiation (254–260 nm) |
| Web of Science | Raeiszadeh and Adeli (2020) A critical review on ultraviolet disinfection systems against COVID-19 outbreak: Applicability, validation, and safety considerations. CS Photonics 2020, 7, 11, 2941–2951 | Not stated | NR | Published | <u>Focus</u><br>To discuss the scientific fundamentals of UV dose requirements for disinfection, protocols for performance validation of UV systems, and safety considerations regarding the use of UV radiation<br><u>Setting</u><br>Experimental and healthcare setting<br><u>Methods of disinfection</u><br>Ultraviolet (UV) disinfection<br><u>Viruses</u><br>SARS-CoV-2, SARS-CoV-1, HCoV strains, MERS-CoV<br><u>Key findings</u><br>Experimental studies on ssRNA viruses, even the early strains of the CoV family, strongly support the opinion that the SARS-CoV-2 can be inactivated by UV radiation. However, the required UV dose and the corresponding level of inactivation | <u>Number of studies</u><br>Not stated<br><u>Quality appraisal</u><br>None<br><u>General comments</u><br>No methods section, no description of study search or selection<br><u>Safety during operation</u><br>One of the most apparent acute effects of UV on the skin is the induction of a cascade of mediators in the skin that together causes “**sunburn**”. UV radiation is also classified as a “**complete carcinogen**”<br>**Ozone generation** is identified among the risks associated with UV disinfection, particularly for air disinfection application |
|  |  |  |  |  | is yet to be determined by the regulatory health organizations for the novel CoV. | In addition to use of appropriate PPE, such as **UV protective goggles and gloves, providing some safety features such as a child lock and motion sensors**, as well as **designing a shield for the UV exposure area** could significantly diminish the chance of human exposure<br><u>Safety and materials</u><br>UV irradiation is known to cause **degradation of materials** that are irradiated (I.e. polymers) |
| VA-ESP | Ramos et al. (2020)<br>Use of ultraviolet-C in environmental sterilization in hospitals: A systematic review on efficacy and safety. Int J Health Sci (Qassim).;14(6):52-65 | 2010 to 2020 | SR | Published | <u>Focus</u><br>To review the literature on the use of ultraviolet-C (UV-C) sterilization to assess its clinical efficacy in reducing risk and transmission of nosocomial infections as well as its associated health safety or hazards<br><u>Setting</u><br>Healthcare facilities<br><u>Methods of disinfection</u><br>Ultraviolet-C<br><u>Bacteria</u><br>Methicillin-resistant Staphylococcus aureus (MRSA), vancomycin-resistant enterococci (VRE), C. difficile<br><u>Virus</u><br>Ebola, influenza, rhinovirus, enterovirus, human metapneumovirus | <u>Number of studies</u><br>Total of full text reviewed (n=17)<br>- Efficacy (n=12)<br>- Safety (n=5)<br><u>Quality appraisal</u><br>A variety of tools were used to assess quality<br><u>General comments</u><br>Only a limited number of studies addressing UV-C safety were included due to the lack of prior research that fit the pre-specified inclusion criteria<br><u>Safety during operation</u><br>Safety study results showed dermal effects of UV-C exposure including DNA lesions, formation of |
|  |  |  |  |  | <u>Key Findings</u><br>The germicidal effect of UV-C is potent against microorganisms including viruses, methicillin-resistant *Staphylococcus aureus*, and vancomycin-resistant enterococci<br>Further studies must still be done to exact a standard for safe exposure dose, especially for 222 nm germicidal lamps. Direct evidence is needed for any targeted implementation of UV-C during Coronavirus Disease 2019 (COVID-19) pandemic | cyclobutane pyrimidine dimers in cells, and effects on the skin's stratum corneum |
| VA-ESP | Kwok et al. (2021)<br>Methods to disinfect and decontaminate SARS-CoV-2: a systematic review of in vitro studies. Ther Adv Infectious Dis. 9:1-12 | Search conducted November 2020 | SR | Published | <u>Focus</u><br>To review cleaning and decontamination methods that have been reported in the literature for SARS-CoV-2<br><u>Setting</u><br>Experimental setting (Laboratory: inoculated SARS-CoV-2 virus onto different types of material including masks, nasopharyngeal swabs, serum, laboratory plates and simulated saliva, tears or nasal fluid)<br><u>Methods of disinfection</u><br>Heat and humidity<br>Ultraviolet (UV) light irradiation*<br>Chemical disinfection<br><u>Viruses</u><br>Coronaviruses, including SARS-CoV-2, SARS-CoV-1, MERS-CoV<br><u>Key findings</u><br>**UV light irradiation:** Different experimental methods testing UV light<br>have shown that it can inactivate the virus. Light of 254–365 nm has been used, including simulated sunlight | <u>Number of studies</u><br>Total of full text reviewed (n=27)<br><u>Quality appraisal</u><br>None<br><u>General comments</u><br>All of the studies took place in laboratory settings rather than real-life clinical settings, so there is insufficient evidence to recommend one type of cleaning procedure over another<br><u>Safety</u><br>No safety issues mentioned with regards to ultraviolet irradiation |
| WHO | Memarzadeh 2021<br>A review of recent evidence for utilizing ultraviolet irradiation technology to disinfect both indoor air and surfaces. Applied Biology. 26(1): 52-6 | Not stated when searches were completed<br>*papers included from 1985 to 2020 | NR | Published | <u>Focus</u><br>To evaluate the applicability, safety and relative contribution of ultraviolet to disinfect air and surfaces in the built environment.<br><u>Setting</u><br>Not explicitly stated. Effective disinfection use mentioned in laboratories, hospitals, clinics, kitchenettes, lobbies, stairwells, animal husbandry areas<br><u>Viruses</u><br>Not specified<br><u>Methods of disinfection</u><br>Ultraviolet germicidal irradiation (UVGI)<br><u>Key findings</u><br>**UV-based sanitizers have the potential for effective application when used in conjunction with other disinfecting means.** The efficacy of UV-based sanitizer technologies are promising but are dependent on numerous environmental, physical and technical factors. UV technologies should not be utilized in isolation and should be considered as an adjunct to protocol-driven standard operating procedures for cleaning and disinfection, hand hygiene practices, and appropriate use of personal protective equipment | <u>Number of studies</u><br>Over 20 source documents (no exact number mentioned)<br><u>Quality appraisal</u><br>None<br><u>General comments</u><br>No methods section, and explicit description how study selection was conducted.<br>**Mentioned lower transmission risk from contact with surfaces containing the virus.** At the time of publication, authors were still awaiting evidence regarding aerosol transmission of SARS-COV-2. They considered SARS-COV-2 as transmitted via virus-laden droplets<br><u>Safety</u><br>UV-A and UV-B wavelengths are harmful to humans and animals because they penetrate the upper layers of skin and can cause skin cancer, cataracts, suppression of the immune system and premature aging of the skin.<br>Ozone may be produced from UV-C lamps emitting UV-C at wavelengths <240nm. |
|  |  |  |  |  |  | UV light disinfection must only take place when rooms are unoccupied to avoid injury to occupants. Other necessary safety measures include warning signage, timers, and motion sensors that shut off the device if any movement is detected inside the room being disinfected |
| VA-ESP | de Oliveria et al. (2020) Disinfectants efficacy and safety for decontamination of surfaces with Covid-19 in healthcare environments: protocol for a systematic review. PROSPERO 2020 CRD42020181294 | Not stated | SR | Protocol | <u>Focus</u><br>Can disinfectants be used to safely and effectively decontaminate surfaces with Covid-19 in healthcare environments?<br><u>Setting</u><br>Healthcare environments<br><u>Methods of disinfection</u><br>Hydrogen peroxide (vapour, liquid, gas plasma), cleaning bleach (sodium hypochlorite), ethanol, isopropyl alcohol, **cleaning with Ultraviolet Light**<br><u>Viruses</u><br>Betacoronavirus. Coronavirus | <u>Quality appraisal</u><br>Quality appraisal will be conducted using GRADE<br><u>General comments</u><br>Potential completion date June 2020<br>Author contacted on 08/09/2021. |

Key: \* RR Rapid review; SR systematic review; NR: narrative review
\*Only key findings related to ultraviolet irradiation extracted

**Table 6:** Summary of secondary research for the potential efficacy of hydrogen peroxide as an air and surface disinfectant against coronaviruses

| Resource | Citation | Recency (Search dates) | Evidence Type* | Status** | Key findings from abstracts | Reviewer comments |
| --- | --- | --- | --- | --- | --- | --- |
| L*OVE<br>PubMed | Kampf et al. (2020a)<br>Persistence of coronaviruses on inanimate surfaces and their inactivation with biocidal agents. *J Hosp Infect.* 104(3): 246–251.<br>Kampf et al. (2020b)<br>Corrigendum to "Persistence of coronaviruses on inanimate surfaces and their inactivation with biocidal agents" [J Hosp Infect 104 (2020) 246-251] *J Hosp Infect.* 105(3):587.<br>Kampf (2020)<br>Potential role of inanimate surfaces for the spread of coronaviruses and their inactivation with disinfectant agents. *Infect Prev Pract.* 2(2): 100044. | 28 January 2020 | SR | Published | <u>Focus</u><br>To review the literature on all available information about the persistence of human and veterinary coronaviruses on inanimate surfaces as well as inactivation strategies with biocidal agents used for chemical disinfection.<br><u>Setting</u><br>Healthcare facilities, although it is unclear if included studies looked at care setting or were experimental<br><u>Methods of disinfection</u><br>Biocidal agents and chemical disinfection*<br><u>Viruses</u><br>Human and veterinary coronaviruses<br><u>Key findings</u><br>MERS and SARS can be efficiently inactivated by 0.5% hydrogen peroxide or 0.1% sodium hypochlorite within 1 minute | <u>Number of studies</u><br>Total of full text reviewed (n=22)<br><u>Quality appraisal</u><br>None<br><u>General comments</u><br>Original review was published first in February 2020. A corrigendum was made in June 2020. The corrigendum refers to the hydrogen peroxide solution mentioned in the review, as the exact composition is unknown. It is assumed that 0.5% hydrogen peroxide is in an accelerated form. (This corrigendum was possibly added as a response to criticism by Lopez Ortega et al. (2020))<br>The paper published in *Infect Prev Pract* is a shorter version of the main article summarised here<br><u>Safety</u><br>No mention of harms, toxicity or safety |
| VA-ESP | Lopez Ortega et al. (2020) | Not stated | Letter to the editor | Published | <u>Focus</u> | <u>Number of studies</u> |
|  | Is 0.5% hydrogen peroxide effective against SARS-CoV-2? Oral Dis 21 June 2020 | *papers included from 2012 to 2020 | (based on SR methodology: protocol registered on PROSPERO (CRD42020190033)) |  | Is 0.5% Hydrogen Peroxide effective against SARS-CoV-2 for surface disinfection?<br><u>Setting</u><br>Dentistry<br><u>Methods of disinfection</u><br>0.5% hydrogen peroxide<br><u>Viruses</u><br>SARS-Cov-2<br><u>Key findings</u><br>Found no study using hydrogen peroxide at 0.5% as a viable substance for surface disinfection<br>The study cited by Kampf does not address the use of 0.5% hydrogen peroxide and there is no study in the literature demonstrating its efficacy as a virucidal agent for surface disinfection either. In fact, we have actually found on PubMed only one study assessing the efficacy of hydrogen peroxide on human coronavirus (SARS), reporting that the virus is inactivated by the substance in the form of vapour at a 35% concentration. | Total of full text reviewed (n=11)<br><u>Quality appraisal</u><br>Meta Analysis of Statistics Assessment and Review Instrument (MAStARI)<br><u>General comments</u><br>This Letter to the editor criticised the review by Kampf et al. (2020a) above |
| VA-ESP | de Oliveira et al. (2020) Disinfectants efficacy and safety for decontamination of surfaces with Covid-19 in healthcare environments: protocol for a systematic review. PROSPERO 2020 CRD42020181294 | Not stated | SR | Protocol | <u>Focus</u><br>Can disinfectants be used to safely and effectively decontaminate surfaces with Covid-19 in healthcare environments?<br><u>Setting</u><br>Healthcare environments<br><u>Methods of disinfection</u><br>Hydrogen peroxide (vapour, liquid, gas plasma), cleaning bleach (sodium<br>hypochlorite), ethanol, isopropyl alcohol, cleaning with Ultraviolet Light | <u>Quality appraisal</u><br>Quality appraisal will be conducted using GRADE<br><u>General comments</u><br>Potential completion date June 2020<br>Author contacted on 08/09/2021. |
|  |  |  |  |  | <u>Viruses</u><br>Betacoronavirus. Coronavirus |  |
| L*OVE | Shimabukuro et al. (2020)<br>Environmental cleaning to prevent COVID-19 infection. A rapid systematic review.<br>San Paulo Med J. 138(6): 505-14 | From 29 April 2020 to 27 May 2020.<br>*papers included from 2000 to 2020 | RR | Published | <u>Focus</u><br>To identify, systematically evaluate and summarize the best available scientific evidence on environmental cleaning to prevent COVID-19 infection.<br><u>Setting</u><br>Experimental and hospital settings<br><u>Methods of disinfection</u><br>70% alcohol, detergent, detergent containing iodine, household bleach, sodium hypochlorite, **hydrogen peroxide**, chlorine dioxide, glutaraldehyde, **ultraviolet irradiation**, plasma air purifier, treating sewage with sodium hypochlorite and chlorine dioxide<br><u>Viruses</u><br>HCoV, SARS-CoV-1, SARS-CoV-2<br><u>Key findings</u><br>Disinfection of environments, especially those in ordinary use, such as bathrooms, needs to be done constantly. **Viral inactivation** was achieved using chlorine-based disinfectants, alcohol, detergents, glutaraldehyde, iodine-containing detergents, **hydrogen peroxide** compounds and household bleaches. Alcohol showed efficient immediate activity | <u>Number of studies</u><br>Total full text reviewed (n=7)<br>- Detergents, bleach, peroxide, aldehydes (n=5)<br>- UV-C ( n=1)<br>- Plasma air purifier (n=1)<br><u>Quality appraisal</u><br>None<br><u>Safety</u><br>Products recommended and authorized by regulatory bodies that presented safety and efficiency with regard to cleaning the environment comprised the intervention. |

**Table 7:** Summary of primary studies for the effectiveness of ozone gas as a decontaminating agent against SARS-CoV 2

| Resource | Citation | Focus | Key findings from abstracts |
| --- | --- | --- | --- |
| **Healthcare environments** |  |  |  |
| Link from<br>Sanitysystem<br>.co.uk<br>(Dublin<br>school ozone<br>machine) | Moccia et al. (2020)<br>Development and improvement of an effective method for air and surfaces disinfection with ozone gas as a decontaminating agent.<br>Medicina (Kaunas). 2020 Nov; 56(11): 578. | <u>Aim</u><br>This study assessed the inactivation of airborne and surface contaminants in healthcare structures using ozone<br><u>Viruses</u><br>Microbial load on surfaces and in the air before and after decontamination<br><u>Setting</u><br>A structured selection of a representative environment of healthcare structures such as high, medium, and low-risk settings in air and examples of hospital furniture | <u>Effectiveness</u><br>The results demonstrated a significant reduction in the microbial count that always fell below the threshold value.<br><u>Safety</u><br>• It was necessary to remark and draw up a precise protocol to be followed by operators;<br>• Use the ozone sanitization cycle only in the absence of people;<br>• Do not use in the presence of flammable substances such as alcohol, petrol, hydrocarbons, bromine, hydrobromic acid, nitrogen oxides and nitroglycerin;<br>• Avoid exposure to UV rays produced by fluorescent lamps;<br>• Seal off the doors and windows of the environments before beginning ozone generation using proper sealing gummed papers in the door and window blows. |

**Table 8:** Summary of secondary research for the effectiveness of no-touch automated disinfection methods against SARS COV-2

| Resource | Citation | Recency<br>(Search dates) | Evidence<br>Type* | Status** | Key findings from abstracts | Reviewer comments |
| --- | --- | --- | --- | --- | --- | --- |
| **Light based technologies** |  |  |  |  |  |  |
| VA-ESP | Cecatto et al (2020)<br>The effectiveness of phototherapy for surface decontamination against viruses. A systematic review.<br>PROSPERO 2020<br>CRD42020184619 | Not reported | SR | Protocol | <u>Focus:</u><br>To investigate the use of light-based therapies (Biophotonics) as antiviral therapy, as well as anti-coronavirus therapy, to discuss the usefulness of biophotonics in surface decontamination against viruses.<br><u>Setting:</u><br>No specific setting reported<br>All surfaces which are contaminated with COVID-19 virus<br><u>Methods of disinfection</u><br>Light-based therapies (Biophotonics)<br><u>Viruses</u><br>Coronavirus | <u>Quality appraisal</u><br>Quality appraisal will be conducted using a variety of validated tools<br><u>General comments</u><br>Potential completion date December 2020<br>Author contacted on 08/09/2021. Author response to full-text request: it has been submitted to a journal and under review. |
| VA-ESP | de Melo Monteiro et al (2020)<br>The COVID-19 outbreak: should dental and medical practices consider UV-C light technology for optimal disinfection on surfaces? A systematic review.<br>PROSPERO 2020<br>CRD42020193961 | Not reported | SR | Protocol | <u>Focus</u><br>Is UV-C light technology efficient for disinfection on surfaces in health care environments?<br><u>Setting</u><br>Dental and medical practices<br><u>Methods of disinfection</u><br>UV-C light technology<br><u>Viruses</u> | <u>Quality appraisal</u><br>Quality appraisal will be conducted using a variety of validated tools<br><u>General comments</u><br>Potential completion date<br>November 2020<br>Author contacted on 08/09/2021.<br>Author response to full-text request: submitted, |
|  |  |  |  |  | Not specified. Subject index include coronavirus, and other communicable diseases | has done revisions, awaiting editor's final decision |
|  | Alvarado-Miranda et al 2020. Analysis of UV technologies for disinfection of public areas: a systematic literature review. IEEE Engineering International Research Conference (EIRCON), 2020, pp. 1-4 | Not stated<br>*papers included from 2012 to 2020 | SR | Published | <u>Focus</u><br>How much is known about the use of UV technology for the disinfection of contaminated areas?<br><u>Setting</u><br>Healthcare setting (39% of included papers) and public areas (21% of included papers)<br><u>Method of disinfection</u><br>UV light technologies<br><u>Viruses</u><br>Not specified<br><u>Key findings</u><br>There are studies on the usefulness of land mobile devices with the use of UV technology to remove and deactivate pathogenic microorganisms from contaminated surfaces in public areas 60%. On the other hand, only 40% of the investigations contemplated in this study did not find sufficient scientific evidence to determine the influence of UV technology on the control of the spread of pathogens in infected areas. | <u>Number of studies</u><br>Total of full text reviewed (n=34)<br>*abstract and PRISMA flowchart shows 34 articles, but in text authors mention 12.<br><u>Quality appraisal</u><br>None<br><u>General comments</u><br>There are issues with the writing style which makes the interpretation of the findings difficult. Some headings, table and figure captions are in Portuguese.<br><u>Safety</u><br>Potential harmful effects to humans mentioned, but not elaborated. UVC was mainly considered safe throughout the paper |
| **Hydrogen Peroxide** |  |  |  |  |  |  |
| Organisation al website<br>PubMed | Tsou et al. (2020) No-touch modalities for disinfecting patient rooms in acute care settings. [Internet]. Rockville (MD): Agency for Healthcare Research and Quality (US); | January 2010 to April 2020 | RR | Published | <u>Focus</u><br>What data exist for the effectiveness of no-touch modalities for disinfecting patient rooms in hospital or acute care settings for:<br>- Respiratory viral pathogens | <u>Number of studies</u><br>Ultra-violet light<br>- Respiratory viruses (1 study)<br>- CDI<br>1 SR, 3 studies)<br>VHP |
|  | 2020 Oct 2. Report No.: 20(21)-EHC021. |  |  |  | - Other pathogens with potential relevance to assessing effectiveness vs. SARS-CoV-2 (specifically CD spores)<br><u>Setting</u><br>Acute settings<br><u>Methods of disinfection</u><br>**Ultraviolet light, vapourous hydrogen peroxide, steam, ozone, chlorine dioxide,** and solid copper surfaces<br><u>Viruses</u><br>Respiratory viruses<br>Clostridioides (formerly Clostridium) difficile (CD) environmental contamination or infection (CDI)<br><u>Key findings</u><br>The effectiveness of no-touch disinfection modalities for disinfecting hospital rooms to decrease respiratory viral infections and CDI remains unclear | - Respiratory viruses (n=0)<br>- CDI 1 SR, 1 study)<br>Solid Cooper surfaces<br>- Respiratory viruses (n=0)<br>- CDI (2studies)<br>Steam, Ozone, Chlorine Dioxide<br>- Respiratory viruses (n=0)<br>- CDI (n=0)<br><u>Quality appraisal</u><br>None<br><u>General comments</u><br>Aside from three studies, the evidence base for no-touch modalities for disinfection of hospital rooms consisted of interrupted time series or single-centre pre/post studies and primarily evaluated impact on CDI rates or room contamination |
| Back chaining | Scarano et al. (2020)<br><u>Environmental Disinfection of a Dental Clinic during the Covid-19 pandemic: A narrative insight.</u> Biomed Res Int . 28;2020:8896812. | inception up to April 30, 2020 | NR | Published | <u>Focus</u><br>To evaluate the scientific literature on the no-touch disinfection procedures in dental clinics aiming to limit transmission via airborne particles or fomites using no-touch procedures for environmental decontamination of dental clinics<br><u>Setting</u><br>Dental clinics | <u>Number of studies</u><br>A total of 86 papers were retrieved by the electronic research. No data on the clinical experience in the decontamination of dental clinics during the pandemic of Covid-19 were detected |
|  |  |  |  |  | <u>Methods of disinfection</u><br>**A**erosolized hydrogen peroxide<br>**H**2O2 vapour<br>**U**ltraviolet C light<br>**P**ulsed xenon<br>**G**aseous ozone<br><u>Viruses</u><br>SARS-Cov-2<br><u>Key Findings</u><br>A general discussion about disinfection and no-touch technologies to improve disinfection of surfaces in dental clinics. In conclusion, manual cleaning and disinfection of environmental surfaces in health care facilities (daily and at patient discharge) are essential elements of infection prevention programs, especially during the SARS-CoV-2 pandemic | <u>Quality appraisal</u><br>Not applicable<br><u>General comments</u> |

**Table 9:** Summary of tertiary research for the effectiveness of SARS CoV-2 no-touch automated disinfection methods

| Resource | Citation | Key Findings |
| --- | --- | --- |
| SAGE | SAGE-EMG 2020a<br><u>Potential application of Air Cleaning devices and personal decontamination to manage transmission of COVID-19</u><br>SAGE minutes 66<br>4<sup>th</sup> November 2020 | What evidence is there that different device technologies may be effective against SARSCoV-2 in terms of air cleaning?<br><u>Portable and fixed room air cleaners designed to be used in occupied spaces</u><br>- As yet little data that demonstrates the effectiveness of most candidate technologies against SARS-CoV-2. Advice in this paper is therefore based on potential effectiveness drawn from the known efficacy of devices against other viruses and the principles of virus transmission.<br>- Air cleaning devices where the primary principle of operation is based on fibrous filtration or germicidal UV (UVC) are likely to be beneficial if deployed correctly (medium confidence). These devices are recommended for settings where the ventilation is poor, and it is not possible to improve it by other means. The efficacy and safety of such devices should be evidenced by relevant test data.<br>- Devices based on other technologies (ionisers, plasma, chemical oxidation, photocatalytic oxidation, electrostatic precipitation) have a limited evidence base that demonstrates effectiveness against SARS-CoV-2 and/or may generate undesirable secondary chemical products that could lead to health effects such as respiratory or skin irritation (medium confidence). These devices are therefore not recommended unless their safety and efficacy can be unequivocally and scientifically demonstrated by relevant test data<br><u>Spray device technologies to decontaminate people in public spaces</u><br>- The use of chemical sprays such as triethylene glycol to clean the air in an occupied space has a limited evidence base in being effective in reducing airborne virus transmission risks, and has potential health effects for those exposed over a long period of time (medium confidence). These approaches are not recommended without further evidence to support their safety and efficacy.<br><u>The use of spray chemicals as a strategy for inactivating virus in the air of occupied rooms</u><br>- Spray booth type devices for decontaminating people are not recommended. They are unlikely to be effective against the virus and have serious health impact and safety concerns. SARSCoV-2 transmission is usually through a result of exhaled virus or via the hands, so even where a person who is infected with COVID-19 has passed through a whole-body disinfection system/device, as soon as they breathe, speak, cough or sneeze they can still spread the virus to others (high confidence)<br>There is good evidence that germicidal UV (GUV) that uses UV-C light and fumigation approaches (particularly Hydrogen Peroxide Vapour (HPV)) are likely to be viable decontamination approaches against SARS-CoV-2 for unoccupied rooms. Both are widely available as commercial systems and are already used in many hospitals for terminal disinfection. UV-C is more challenging to apply well in a complex space with surfaces in shadow but 'shadowing' effects can also affect fumigation efficacy, with areas facing away from delivery equipment or positions on the underside of room surfaces the most challenging to reach |
| SAGE | SAGE-EMG 2020b<br>Application of UV disinfection, visible light, local air filtration and fumigation technologies to microbial control.<br>SAGE minutes 37<br>19<sup>th</sup> May 2020 | This paper summarises evidence for ultraviolet (UV) disinfection, visible light, local air filtration and fumigation technologies to be applied to control COVID-19 transmission<br>- There is good evidence that **germicidal UV (GUV)** that uses **UV-C light** and **fumigation approaches** (particularly **Hydrogen Peroxide Vapour (HPV)**) are likely to be viable decontamination approaches against SARS-CoV-2 for unoccupied rooms.<br>- Both UV-C and fumigation decontamination require a sufficient duration of exposure to be effective. As such they are more likely to be effective as part of a terminal cleaning process rather than daily disinfection. This is particularly the case for fumigation which requires 30-90min cycle time, plus time for aeration to remove of any excess fumigants. UV carousel devices are typically deployed for between 20 and 45 minutes, depending on the room to be treated, but may also require moving and repeat treatment to overcome shadowing effects.<br>- Removal of fumigant by aeration is a particular concern for fumigation approaches that should be considered particularly in environments with a high level of soft furnishings<br>- Both UV-C and fumigation decontamination approaches have significant safety considerations and should only be carried out by trained staff with appropriate risk assessments and controls in place.<br>- Upper room GUV has significant safety considerations which must be taken into consideration in the design, installation and operation |
| TAG-E | TAG-E 2021<br>Air cleaning devices<br>29 January 2021 | Informed by the two above reports from SAGE-EMS<br><u>Key Findings</u><br>- Air cleaning devices are not a substitute for ventilation and every effort should be made to increase ventilation before considering them (high confidence).<br>- Air cleaning devices may be of benefit in poorly ventilated spaces where it is not possible to improve it by other means (medium confidence) but are of little use in well ventilated spaces.<br>- Air cleaning devices where the primary principle of operation is based on fibrous filtration (such as HEPA filters) and germicidal UV (UVC) are likely to be beneficial if deployed correctly (medium confidence).<br>- The performance of most devices is based on data measured in idealised controlled environments and is likely to be different and often lower in a real-world setting (high confidence). Caution is therefore advised when considering manufacturer's performance data.<br>- SAGE suggested that further research is needed on the efficacy of devices including evidence of the technology against SARS-CoV-2 virus (or a suitable viral surrogate) and other pathogens and their performance in real-world settings. For them to be effective, there needs to be sufficient time for the air in the room to pass through the purifier.<br>There may be unintended consequences from the application of air cleaning devices |
|  | US EPA (2020)<br>Ozone generators that are sold as air | - Much of the written material on the use of ozone indoors makes claims or conclusions without substantiation and sound science. Only peer-reviewed, scientifically supported findings and conclusions were relied upon in developing this document. |
|  | cleaners.<br>https://www.epa.gov/in-door-air-quality-iaq/ozone-generators-are-sold-air-cleaners | - No agency of the federal government has approved these devices for use in occupied spaces.<br>- Whether in its pure form or mixed with other chemicals, ozone can be harmful to health.<br>- The same chemical properties that allow high concentrations of ozone to react with organic material outside the body give it the ability to react with similar organic material that makes up the body, and potentially cause harmful health consequences.<br>- Damage to the lungs, decrease in lung function, inflammation of lung tissue, chest pain, coughing, shortness of breath, throat irritation, aggravation of asthma and higher susceptibility to respiratory infection are all listed as health effects of ozone.<br>- Recovery from the harmful effects can occur following short-term exposure to low levels of ozone, but health effects may become more damaging and recovery less certain at higher levels or from longer exposures<br>- The phrase "good up high - bad nearby" has been used by the U.S. Environmental Protection Agency (EPA) to make the distinction between ozone in the upper and lower atmosphere.<br>- Available scientific evidence shows that at concentrations that do not exceed public health standards, ozone has little potential to remove indoor air contaminants, viruses, bacteria, mould or other biological pollutants.<br>- Ozone concentrations would have to be 5 - 10 times higher than public health standards allow before the ozone could decontaminate the air sufficiently to prevent survival and regeneration of the organisms once the ozone is removed.<br>- Even with high levels of ozone, contaminants embedded in porous material may not be affected at all.<br>- High concentrations of ozone in air, when people are not present, are sometimes used to help decontaminate an unoccupied space from certain chemical or biological contaminants or odours, however little is known about the chemical by-products left behind by these processes, which of themselves can be very reactive and capable of producing irritating and corrosive by-products.<br>- While high concentrations of ozone in air may sometimes be appropriate in these circumstances, conditions should be sufficiently controlled to insure that no person or pet becomes exposed.<br>- For many of the chemicals with which ozone does readily react, the reaction can form a variety of harmful or irritating by-products<br>- In addition to aldehydes, ozone may also increase indoor concentrations of formic acid, both of which can irritate the lungs if produced in sufficient amounts.<br>- The actual concentration of ozone produced by an ozone generator depends on many factors including, the area of the room, ventilation (doors being open), the number of materials and furnishings that can absorb or react with the ozone.<br>- Ozone can adversely affect indoor plants, and damage materials such as rubber, electrical wire coatings and fabrics and artwork containing susceptible dyes and pigments. |

Key: SAGE-EMG: Scientific Advisory Group for Emergencies- Environmental and Modelling Group; WG-TAG: Welsh Government-Technical Advisory Environmental Sub Group

**Table 10.** Table of potential primary research studies

| Primary Research |  |  |  |
| --- | --- | --- | --- |
| Aspect of Question explored | Study type | Total | Other Comments |
| Effectiveness of ozone machines on other viruses and bacterium | Primary study | 5 | Yano et al. (2020): Letter to the editor with regard to a study about Inactivation of severe acute respiratory syndrome coronavirus 2 (SARS-CoV-2) by gaseous ozone treatment.<br>Sharma and Hudson 2008: Evaluated the efficacy of a portable ozone-generating machine, equipped with a catalytic converter and an accessory humidifier, to inactivate 15 different species of medically important bacteria.<br>Moat et al. 2009: Effectiveness of a novel ozone-based system for the rapid high-level disinfection of health care spaces and surfaces (ozone and hydrogen peroxide vapour)<br>Zoutman et al. 2011: Effectiveness of a novel ozone-based system for the rapid high-level disinfection of health care spaces and surfaces<br>Tseng et al 2006: The effects of ozone concentration, contact time, different capsid architecture of virus and relative humidity on inactivating airborne viruses by ozone were evaluated in a laboratory test chamber.<br>Tarka and Nitsch-Ousch 2020: Peroxone vapour, a combination of hydrogen peroxide and ozone in the decontamination of 50 surfaces in 10 hospital rooms. |
| No touch technologies | Primary study | 1 | Haydar et al. 2021: A high-level disinfection cabinet, electrostatic sprayer and an ultraviolet-C light box on a variety of viruses and bacterium for toys in a paediatric ward |

## Acknowledgements

The authors would like to thank Zakhyia Begum, Arwyn Edwards and Miranda Morton for their contributions during stakeholder meetings to guide the focus of the evidence summary and interpret findings.

## About the Wales COVID-19 Evidence Centre

The WC19EC integrates with worldwide efforts to synthesise and mobilise knowledge from research.

We operate with a core team as part of Health and Care Research Wales, are hosted in the [Wales Centre for Primary and Emergency Care Research (PRIME)](http://www.primecentre.wales/), and are led by [Professor Adrian Edwards of Cardiff University](https://www.cardiff.ac.uk/people/view/123022-edwards-adrian).

The core team of the centre works closely with collaborating partners in [Health Technology Wales](https://www.healthtechnology.wales/), Wales Centre for Evidence-Based Care, [Specialist Unit for Review](https://www.cardiff.ac.uk/specialist-unit-for-review-evidence), [Evidence centre](https://www.cardiff.ac.uk/specialist-unit-for-review-evidence), [SAIL Databank](https://healthandcareresearchwales.org/about-research-community/secure-anonymised-information-linkage-sail-databank), [Bangor Institute for Health & Medical Research/Health and Care Economics Cymru](https://www.bangor.ac.uk/health-sciences/research/index.php.en), and the [Public Health Wales Observatory](https://phw.nhs.wales/services-and-teams/observatory/).

Together we aim to provide around 50 reviews per year, answering the priority questions for policy and practice in Wales as we meet the demands of the pandemic and its impacts.

### Director

Professor Adrian Edwards

### Contact Email

**Website:** https://healthandcareresearchwales.org/about-research-community/wales-covid-19-evidence-centre

## APPENDIX – Resources searched

11. APPENDIX – Resources searched
| Resource |  | Success or relevancy of the retrieval |
| --- | --- | --- |
| **Priority COVID resources for reviews** |  |  |
| Cochrane COVID Review Bank | Scrolled through list for relevant reviews | Searched, nothing found |
| VA-ESP | Keyword search<sup>a</sup><br>or inactivation, hydrogen peroxide, ultraviolet, UV-C | Searched, results found 66 results, **10** potentially relevant |
| L*OVE – COVID-19 | Ozone<br>Keyword search<sup>b</sup><br>or inactivation, hydrogen peroxide, ultraviolet, UV-C | Searched, results found 4 results, 0 relevant<br>65 results, **10** potentially relevant |
| Collabovid |  | Not searched, maybe relevant |
| **Additional COVID resources for reviews** |  |  |
| LitCovid | Keyword search<sup>d</sup> | Searched, results found 599 results, **6** potentially relevant |
| EPPI Centre: Living map of systematic reviews of social science research on COVID-19 | Keyword search<sup>a</sup> | Searched, nothing found |
| EPPI Centre – Living map of COVID-19 research | Keyword search<sup>a</sup> | Searched, nothing found |
| **For technology / treatment questions** |  |  |
| International HTA database (ITS-HTA) | Keyword search<sup>b</sup><br>Keyword search<sup>c</sup><br>Ozone | Searched, results found 0 results<br>0 results<br>1 result, 0 relevant |
| EUnetHTA – COVID 19 response |  | Not searched, not relevant |
| **Additional COVID resources for primary studies** |  |  |
| L*OVE primary studies |  | Not searched, maybe relevant |
| Cochrane COVID-19 Study Register |  | Not searched, maybe relevant |
| LitCovid |  | Not searched, maybe relevant |
| **Secondary research resources for reviews (non-COVID-19)** |  |  |
| Trip | Keyword search<sup>e</sup> | Searched, results found<br>8 results, **3** potentially relevant |
| [Cochrane Database of Systematic Reviews (CDSR)](https://www.cochranelibrary.com/cdsr/reviews)<br>https://www.cochranelibrary.com/cdsr/reviews | Keyword search<sup>b</sup> | Searched, nothing found |
| Campbell Collaboration |  | Not searched, not relevant |
| JB I (via OVID) |  | Not searched, not relevant |
| [Epistemonikos](https://www.epistemonikos.org/en/advanced_search)<br>https://www.epistemonikos.org/en/advanced_search |  | Not searched, maybe relevant |
| PROSPERO |  | Not searched, not relevant |
| Pubmed Clinical Queries |  | Not searched, maybe relevant |
| PubMed | Ozone AND corona* OR corono* OR COVID*OR SARS-C*)<br>(clean* OR disfect* OR decontamin* OR santif* OR eliminat* OR inactivat* AND (corona* OR corono* OR COVID*OR SARS-C*))<br>“Hydrogen peroxide” AND (corona* OR corono* OR COVID*OR SARS-C*)<br>Ultraviolet or uv* AND (corona* OR corono* OR COVID*OR SARS-C*)<br>“automated room” or “no touch” or no-touch AND | Searched, results found<br>3 results, 0 relevant<br>44 results, **7** potentially relevant<br>4 results, 0 relevant<br>18 results, **2** potentially relevant<br>26 results, **6** potentially relevant |
|  | clean* OR disfect* OR<br>decontamin* OR santif* OR<br>eliminat* OR inactivat |  |
| Medline<br>Embase<br>Web of Science<br>Imported into endnote, duplicates and removed and searched within titles for the word review | Full searches available on<br>request<br>Ozone or air AND COVID<br>AND disinfection or<br>decontamination | Searched, results found<br>31 results, 7 potentially relevant |
| Scopus<br>Embase<br>Web of Science<br>Imported into endnote, duplicates and removed and searched within titles for the word review | Full searches available on<br>request<br>Ozone AND harms | Searched, results found<br>72 results, 7 potentially useful |
| Health & Environmental Research Online (HERO) | Keyword search<sup>a</sup> | Searched, results found<br>20 results, 6 potentially relevant |
| **Secondary resources for reviews relevant to local/UK context** |  |  |
| Public Health England (PHE) COVID-19 Rapid Reviews | Scrolled through list for<br>relevant reviews | Searched, nothing found |
| NICE resources for COVID reviews | Scrolled through list for<br>relevant reviews | Searched, nothing found |
| Healthcare Improvement Scotland – COVID-19: Evidence for Scotland | Scrolled through list for<br>relevant reviews | Searched, nothing found |
| Ireland, HSE Library, Covid-19 Summaries of Evidence | Scrolled through list for<br>relevant reviews | Searched, results found<br>1 potentially relevant |
| HIQA Health Information and Quality Authority (Ireland) – Rapid reviews | Keyword search<sup>a</sup> | Searched, nothing found |
| SAGE | Keyword search<sup>a</sup> | Searched, results found<br>2 potentially relevant |
| Wales: TAG | Keyword search<sup>a</sup> | Searched, results found<br>1 potentially relevant |
| **Secondary resources for reviews produced by key international organisations** |  |  |
| ECDC European Centre for Disease Prevention and Control (COVID-19 outputs) | Keyword search<sup>a</sup> | Searched, results found<br>1 potentially relevant |
| CDC centre for Disease Control and Prevention<br>Science Briefs (US) | Keyword search<sup>a</sup> | Searched, results found<br>1 potentially relevant |
| AHRQ Agency for Healthcare Research and Quality (US) | Keyword search<sup>a</sup> | Searched, results found<br>1 potentially relevant |
| NASEM The National Academy of Sciences Engineering Medicine | Keyword search<sup>a</sup>: | Searched, nothing found |
| Coronavirus Resources Collection (US) |  |  |
| Australian National COVID-19 Clinical Evidence Task Force - Living Guidelines |  | Not searched, not relevant |
| NCCMT COVID-19 rapid reviews (Canada) | Scrolled through list for relevant reviews | Searched, results found **1** potentially relevant |
| WHO Global literature on coronavirus disease (COVID-19) database | Surface* AND disinfect* or decontamination or ozone AND review | Searched, results found 11 results, **5** potentially relevant |

Keyword search<sup>a</sup>: Surface; Ozone; Disinfection; Decontamination
Keyword search<sup>b</sup>: surface\* or room\* AND (clean\* or disfect\* or decontamin\* or santif\* or remov\* or mitigat\* or eliminat\* or reduc\*)
Keyword search<sup>c</sup>: ozone AND (clean\* or disfect\* or decontamin\* or santif\* or remov\* or mitigat\* or eliminat\* or reduc\*)
Keyword search<sup>d</sup>: Surface\* AND disinfect\* / Surface\* AND decontamin\* / ozone AND disinfection / ozone AND decontamination / ozone AND generator\* / ozone AND machine\* / ozone AND device\* / gaseous ozone / air AND clean AND device / air AND clean AND technology / air AND decontamination / "air disinfection" / "air purification" / "air sterilisation" / "air sterilization" / "room decontamination" / "viral deactivation"
Keyword search<sup>e</sup>: surface\* or room\* AND (clean\* or disfect\* or decontamin\* or santif\* or remov\* or mitigat\* or eliminat\* or reduc\*)

